# Psychological Resilience in Adolescence as a function of Genetic Risk for Major Depressive Disorder and Alzheimer’s Disease

**DOI:** 10.1101/2022.07.19.22277815

**Authors:** Raluca Petrican, Alex Fornito

**Affiliations:** Cardiff University Brain Research Imaging Centre (CUBRIC), School of Psychology, Cardiff University, Maindy Road, Cardiff, CF24 4HQ, United Kingdom; The Turner Institute for Brain and Mental Health, School of Psychological Sciences, and Monash Biomedical Imaging, Monash University, Melbourne, VIC, Australia

**Keywords:** Psychological Resilience, Major Depressive Disorder, Alzheimer’s Disease, Adolescent Development, Polygenic Risk, Transcriptomics

## Abstract

Major Depressive Disorder (MDD) and Alzheimer’s Disease (AD) are two pathologies linked to prior stress exposure and altered neurodevelopmental trajectories. As a putative antecedent to AD, MDD could be key to understanding the neurobiological changes that precede the clinical onset of AD by decades. To test this hypothesis, we used longitudinal data from the Adolescent Brain and Cognitive Development study (N_total_ = 980, 470 females) and investigated overlapping connectomic, transcriptomic, and chemoarchitectural correlates of adjustment to stressors (i.e., resilience) among adolescents at genetic risk for AD and MDD, respectively. The potential for perinatal adversity to directly and/or indirectly, via accelerated biological ageing, foster resilience (i.e., “inoculation” effects) was also probed. We identified two distinguishable neurodevelopmental profiles predictive of resilience among MDD-vulnerable adolescents. One profile, expressed among the fastest developing youth, overlapped with areas of greater dopamine receptor density and reflected the maturational refinement of the inhibitory control architecture. The second profile distinguished resilient MDD-prone youth from psychologically vulnerable adolescents genetically predisposed towards AD. This profile, associated with elevated GABA, relative to glutamate, receptor density, captured the longitudinal refinement and increasing context specificity of incentive-related brain activations. Its transcriptomic signature implied that poorer resilience among AD-prone youth may be associated with greater expression of MDD-relevant genes. Our findings are compatible with the proposed role of MDD as a precursor to AD and underscore the pivotal contribution of incentive processing to this relationship. They further speak to the key neuromodulatory role of DA-gonadal hormone interactions in fostering resilience in adolescence.

**Significance Statement:** Environmental stressors can substantially alter brain maturation and incur lifelong costs. Using longitudinal data, we characterise two developmental profiles correlated with positive adjustment to environmental challenges (i.e., resilience) among adolescents at genetic risk for two stress-related conditions, Alzheimer’s Disease (AD) and Major Depressive Disorder (MDD), respectively. One dopamine-related profile typified the fastest developing MDD-prone adolescents and reflected the neural maturation of the inhibitory control architecture. The second profile, neurochemically linked to excitation/inhibition balance, indicated the developmental refinement of motivational pathways, distinguishing resilient MDD-prone from psychologically vulnerable AD-prone teens. Its transcriptomic signature supported the posited role of MDD as an antecedent to AD. Our results unveil candidate neurobiological mechanisms supporting lifespan resilience against both psychiatric and neurological conditions linked to stress exposure.

## Background

Adolescence is a critical developmental phase, marked by a multitude of interdependent neurobiological and socio-environmental changes that are programmed to maximise adjustment to one’s milieu (1, 2). Unsurprisingly, this is also the life stage in which more than a third of all diagnosed mental disorders have their onset (3), a finding that underscores the urgency in comprehensively characterising the developmental markers of subsequent adaptation versus vulnerability (4, 5).

Alzheimer’s Disease (AD) and Major Depressive Disorder (MDD) are two conditions typified by altered brain maturation/ageing trajectories, in part stemming from systemic inflammation processes (6–8). Robustly linked to greater stress susceptibility and exposure, as well as genes regulating lifespan neurodevelopment, the two disorders have been posited to share a causal connection, with MDD and broader predispositions towards experiencing negative affect being regarded as risk factors or even early symptoms of AD (9–14).

Although the clinical onset of AD occurs mid- to late adulthood, compelling evidence suggests that the preceding decades are marked by significant cellular changes linked to more subtle cognitive-behavioural markers (15). Given that AD-relevant genetic risk factors are under considerable environmental modulation (16), identification of early life neurodevelopmental alterations among vulnerable individuals could be instrumental in designing interventions to slow down or even prevent progression towards dementia.

Leveraging the purported relationships among stress, MDD and AD, we used a longitudinal design to probe the overlap of the macroscale (functional connectomic) and molecular (neurochemical and transcriptomic) brain correlates of psychological resilience (i.e., positive adjustment to environmental challenges, [17, 18]) as a function of genetic risk for MDD versus AD. Overlapping substrates could unveil common pathways linking stress exposure to both AD and MDD, thereby justifying subsequent investigations into whether neurodevelopmental mechanisms that promote early life resilience may also foster resistance to AD in older adulthood.

Secondarily, we sought to extend findings on the lifelong neurodevelopmental consequences of perinatal adversity exposure by characterising its potential risk-enhancing or protective (“inoculation”-like) role with regards to resilience, whether manifest directly or indirectly via an altered rate of biological ageing (i.e., pubertal timing) (cf. 19-25). Our complete model is depicted in Figure 1.

**Figure 1.**
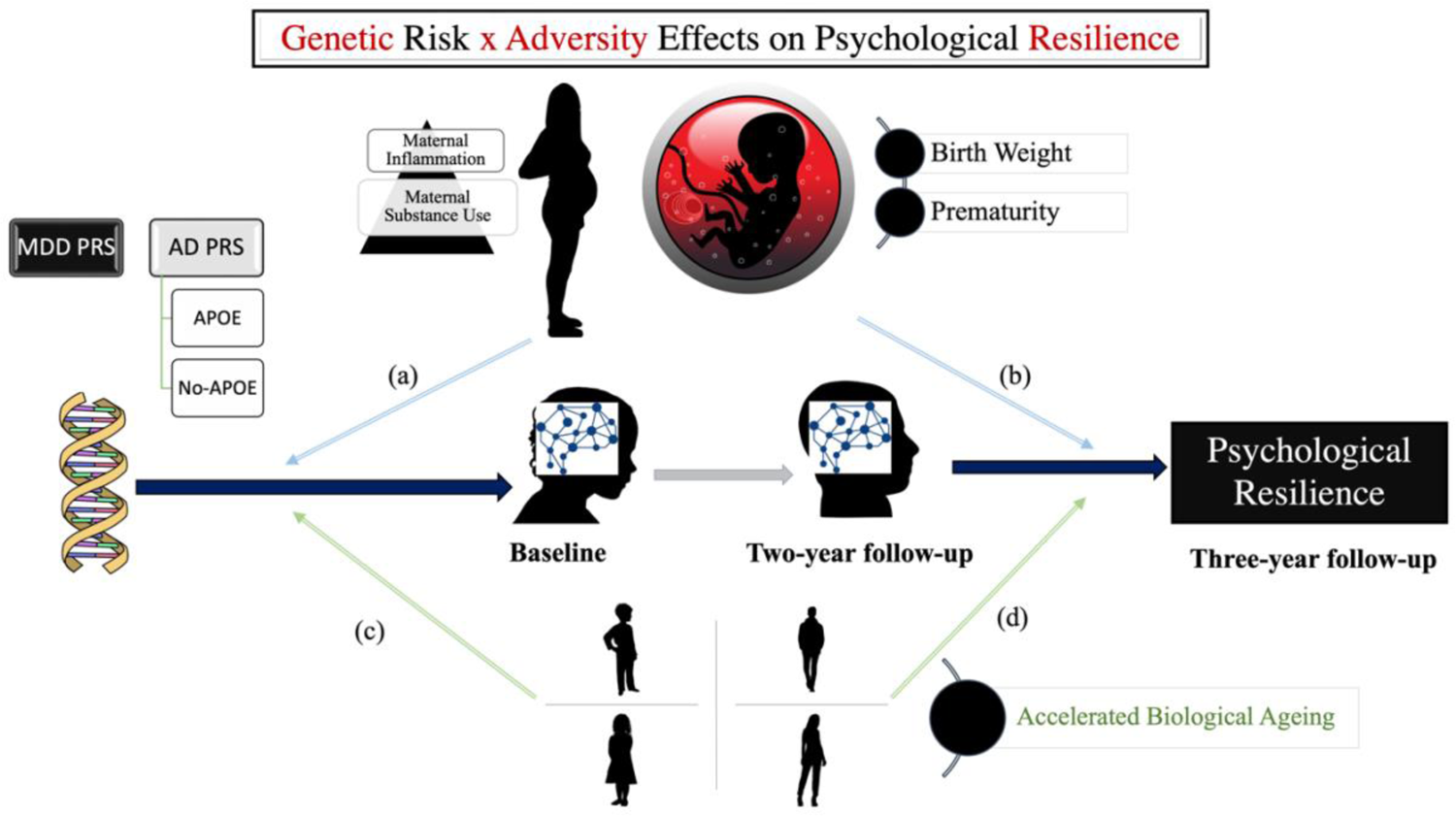
Schematic representation of the conceptual/measurement model. Genetic risk for AD (APOE vs no-APOE-based, see Method for details) and MDD, respectively, were predicted to impact functional brain maturation, specifically, longitudinal fine-tuning and contemporaneous, cross-context differentiation, as observed on inhibitory control and incentive processing tasks. The primary goal was to identify neurodevelopmental patterns that would alleviate the expected negative impact of AD/MDD genetic risk on resilience. The moderating, potentially protective (“inoculation”-like effect) effects of perinatal adversity and accelerated biological ageing on gene-brain (paths a, c) and brain-resilience (paths b, d) were also probed.

In light of its well-demonstrated utility in modelling trajectories of both normative brain maturation and (transdiagnostic) pathology, including AD, a network neuroscience approach was applied to longitudinal multimodal data from the Adolescent Brain and Cognitive Development (ABCD) study (26–28). Genetic vulnerability to AD and MDD, respectively, was quantified using polygenic risk scores (PRS, [29]) derived from genome-wide association studies (GWAS) (30–31). Developmental connectomic correlates of resilience were estimated in reference to two mental processes, inhibitory control and reward processing, which change substantially in adolescence, are highly susceptible to adversity and predictive of subsequent (psycho)pathology, including later life vulnerability to dementia (32–38). Molecular correlates of resilience were quantified using group-based normative maps of gene expression and receptor density focused on four neutransmitters (dopamine [DA], serotonin [5-HT], glutamate [GLU], gamma-aminobutyric acid [GABA]) implicated in cognitive control, incentive processing, susceptibility to stress/resilience, as well as AD/MDD vulnerability, more specifically (39–49).

## Results

### Mesoscale Functional Brain Correlates of Resilience and AD/MDD Risk

To characterise functional connectomic features linked to resilience, we applied a network-based clustering technique, referred to as “multilayer community detection” (50) to fMRI data collected during two tasks assessing inhitory control and incentive processing, respectively (26). Using the Schaefer 300 parcel-functional atlas (51) and the “flexibility” function from the Network Community Toolbox (52), we estimated longitudinal (baseline-to-2-year follow-up) within-task and cross-task (inhihibitory control/incentive processing) contemporaneous differences in functional brain architecture (i.e., changes in each parcel’s functional community assignment [53]). The first index gauged fine-tuning of task-specific functional brain architecture. The second index reflected context-specificity in neural coding (i.e., inhibitory control-incentive processing differentiation), a cross-species marker of adaptive functioning and resilience (54–56). Partial least squares analysis (PLS) was subsequently applied to parcel-specific “flexibility” indices to identify connectomic features linked to resilience and/or genetic risk for AD/MDD.

PLS analysis 1 identified two latent variables (LVs, *p*-values of .0002 and .001, respectively), which accounted for 18.20% and 12.83%, respectively, of the brain-behaviour covariance. The first extracted LV related greater MDD risk, accelerated biological ageing and resilience to longitudinal increases in Control, somatomotor (SM), Salience/Ventral Attention (SAL/VAN), default mode (DMN) and visual (VIS) functional reconfiguration on the inhibitory control task (see Figure 2-a, b). The second LV linked VIS-related longitudinal and cross-task functional reorganisation at follow-up to higher resilience in the presence of stronger MDD, but weaker AD, risk (see Figure 3-a, b). Specifically, it implied that psychological health may hinge on refinement of VIS responses to incentives and rising task-specificity in processing.

**Figure 2.**
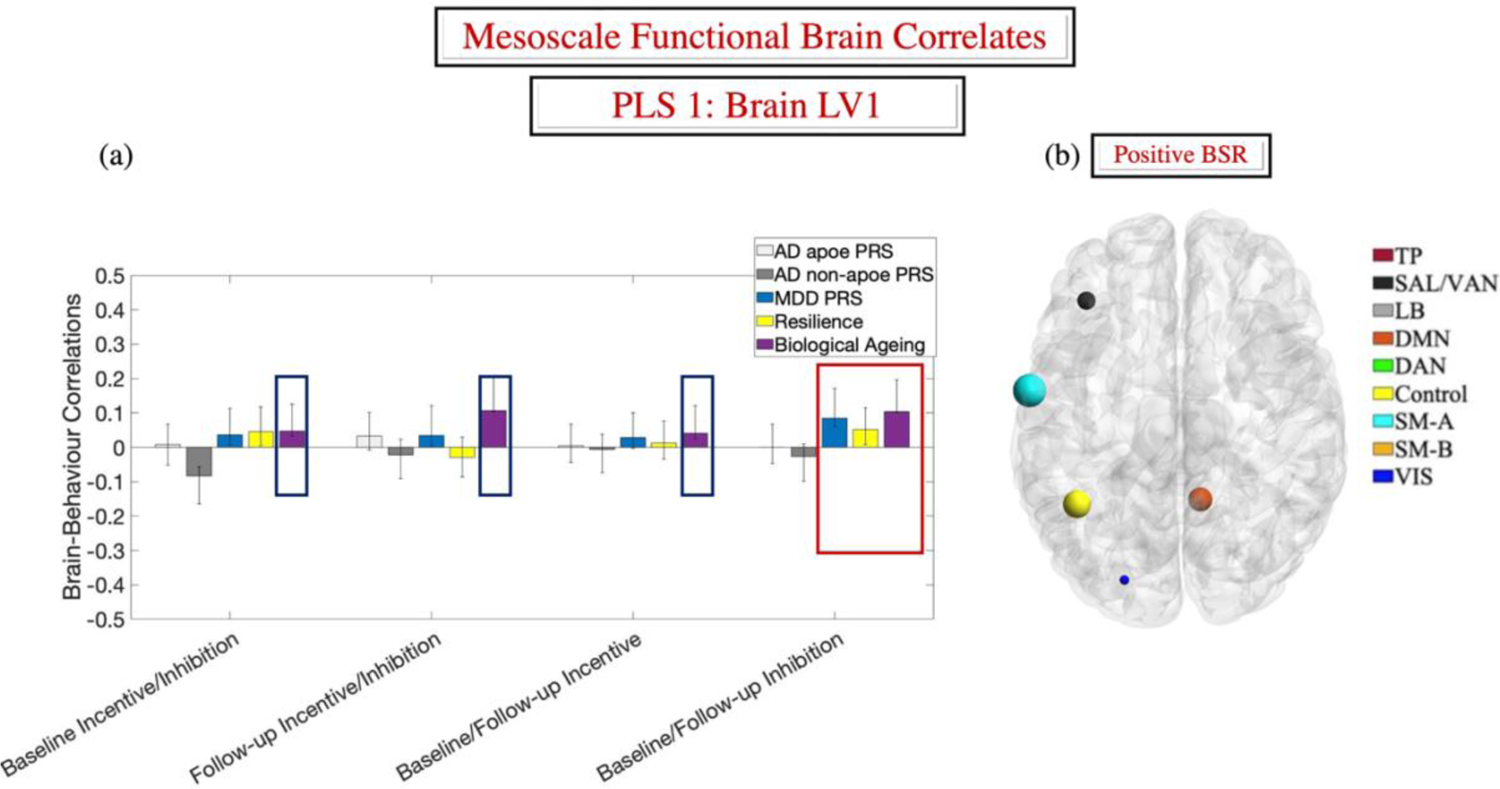
First extracted LV from the behavioral-PLS analysis linking AD/MDD genetic risk, resilience, and biological ageing to longitudinal and contemporaneous cross-context differentiation of the functional brain architecture relevant to incentive processing vs inhibitory control. Panel (a) shows the correlations between the behavioral variables and the brain scores in each condition. Panel (b) depicts the Schaefer parcels with robust loadings (absolute value BSR > 3) on the LV in panel (a) and visualized with the BrainNet Viewer (http://www.nitrc.org/projects/bnv/) (Xia et al., 2013). Parcel colours reflect Schaefer et al.’s network assignments. In panel (b), the size of the parcels is proportional to their associated absolute value BSR. Error bars are the 95% confidence intervals from the bootstrap procedure. Confidence intervals that do not include zero reflect robust correlations between the respective behavioral variable and the brain score in a given condition across all participants. In panel (a), hypothesis-relevant, significant brain-behavior correlations, replicated across all the supplemental analyses, are within red line rectangles. Significant brain-behavior correlations unrelated to our hypotheses are in blue rectangles. All the reported PLS analyses had been conducted on parcel-specific indices. Network labels attached to the parcels are included for informational purposes only. LV= latent variable. PRS = polygenic risk score. BSR = bootstrap ratio. Schaefer networks: TP= temporo-parietal. SAL-VAN = salience/ventral attention. LB = limbic. DMN = default mode. DAN = dorsal attention. SM-A =somatomotor-A. SM-B =somatomotor-B. VIS = visual.

**Figure 3.**
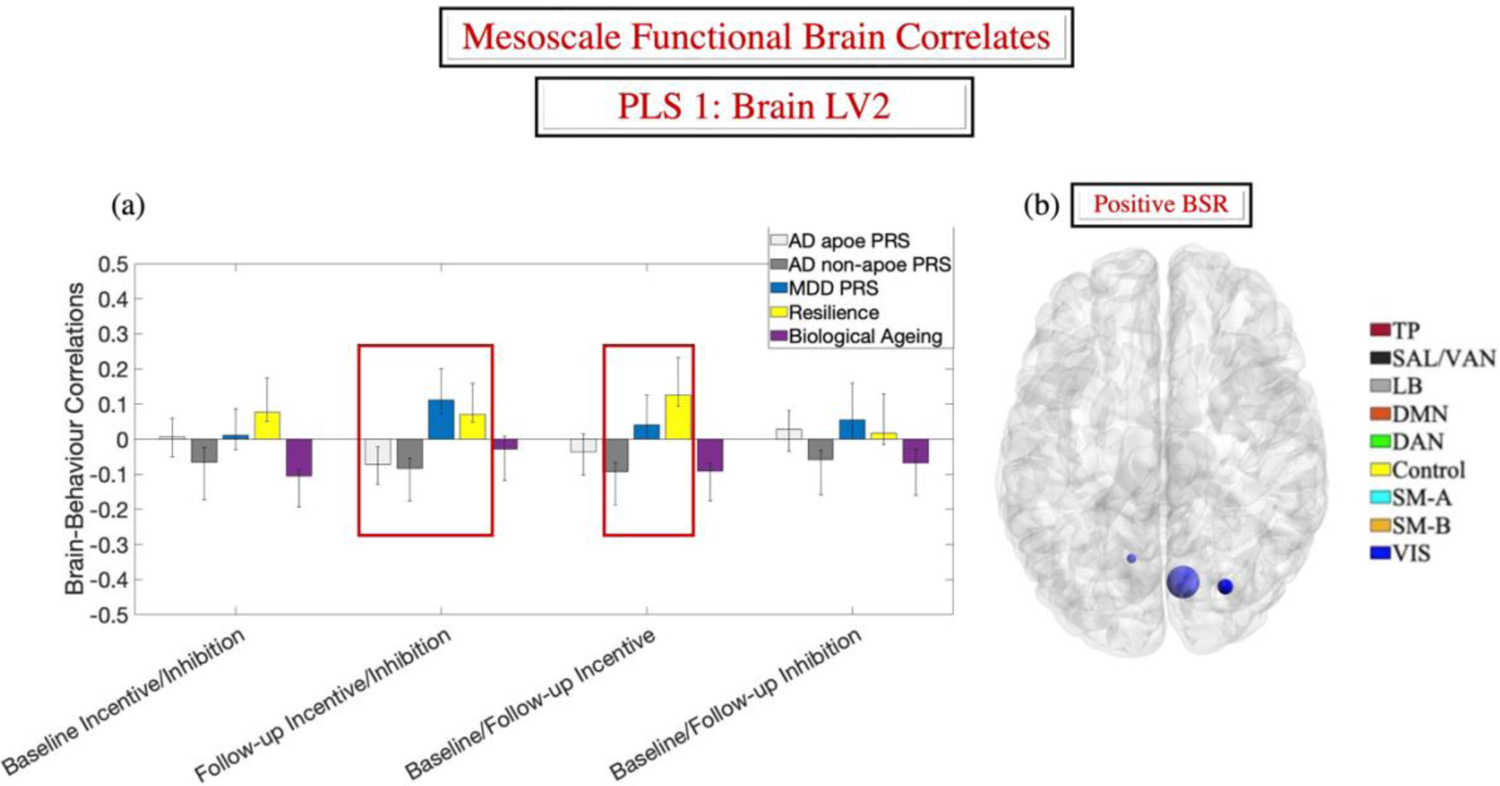
Second extracted LV from the behavioral-PLS analysis linking AD/MDD genetic risk, resilience, and biological ageing to longitudinal and contemporaneous cross-context differentiation of the functional brain architecture relevant to incentive processing vs inhibitory control. Panel (a) shows the correlations between the behavioral variables and the brain scores in each condition. Panel (b) depicts the Schaefer parcels with robust loadings (absolute value BSR > 3) on the LV in panel (a) and visualized with the BrainNet Viewer (http://www.nitrc.org/projects/bnv/) (Xia et al., 2013). Parcel colours reflect Schaefer et al.’s network assignments. In panel (b), the size of the parcels is proportional to their associated absolute value BSR. Error bars are the 95% confidence intervals from the bootstrap procedure. Confidence intervals that do not include zero reflect robust correlations between the respective behavioral variable and the brain score in a given condition across all participants. In panel (a), hypothesis-relevant, significant brain-behavior correlations replicated across all the supplemental analyses, are within red line rectangles. Significant brain-behavior correlations unrelated to our hypotheses are in blue rectangles. All the reported PLS analyses had been conducted on parcel-specific indices. Network labels attached to the parcels are included for informational purposes only. LV= latent variable. PRS = polygenic risk score. BSR = bootstrap ratio. Schaefer networks: TP= temporo-parietal. SAL-VAN = salience/ventral attention. LB = limbic. DMN = default mode. DAN = dorsal attention. SM-A =somatomotor-A. SM-B =somatomotor-B. VIS = visual.

The above PLS results were replicated with a distinct functional brain atlas (see Supplemental Materials). Exploratory PLS analyses conducted with males and females as two separate groups did not reveal any reliable sex differences with regards to the connectomic correlates of resilience or AD/MDD genetic vulnerability.

### Microscale Correlates of Resilience and AD/MDD Risk

To characterise the transcriptomic signature of the two brain LVs described above, we used the abagen toolbox (57) and quantified gene expression levels in each of the Schaefer parcels (see *Methods*). Two PLS analyses were subsequently conducted on the resilience-relevant brain LVs and the parcel x gene matrices outputted by the abagen toolbox.

### Gene-brain PLS 1: Left-hemisphere only

The gene-brain PLS analysis identified a sole LV (*p* = .017), which accounted for 70% of the brain-gene covariance. The extracted gene LV was positively correlated with both the brain LV1 (cf. Figure 2), *r* = .32, 95% CI = [.25; .45] and brain LV2 (cf. Figure 3), *r* = .48, 95% CI = [.40; .62].

### Gene-brain PLS 2: Bi-hemispheric

The above brain-gene pattern was replicated using the bi-hemispheric gene expression data from the Schaefer atlas (*p* = .017, 63% brain-gene covariance explained; see Figure 4-a, b for the spatial expression profile of the gene LV). As expected, the identified gene LV was robustly related to both brain LVs (LV1: *r* = .30, 95%CI = [.24; .39]; LV2: *r* = .42, 95%CI = [.36; .51]).

**Figure 4.**
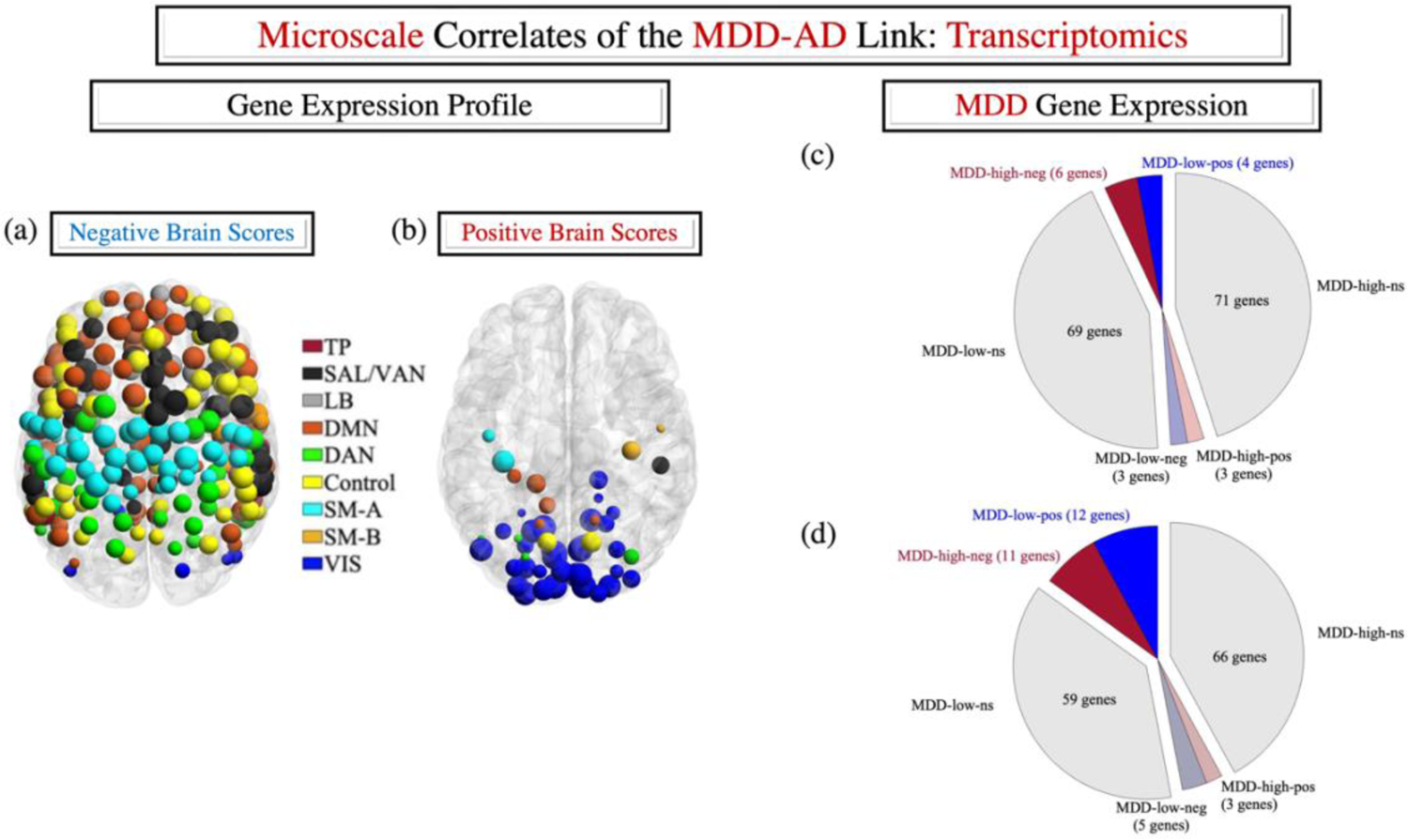
Transcriptomic correlates of the AD-MDD link. Panels (a) and (b) represent the spatial expression map of the gene LV identified in the gene-brain behavioral PLS analysis. The parcels were visualized with the BrainNet Viewer (http://www.nitrc.org/projects/bnv/) (Xia et al., 2013). Parcel colours reflect Schaefer et al.’s network assignments and their size is proportional to how strongly they express the gene LV (i.e., the absolute value of the associated brain score). Panels (c) and (d) depict the results of the MDD/no-APOE AD PRS overlap analyses, using only left-hemisphere (c) or bi-hemispheric (d) data. As described in the text, these are based on the gene LV depicted in panels (a) and (b), which was negatively correlated with the no-APOE PRS (see Figure 3-a). LV= latent variable. BSR = bootstrap ratio. Schaefer networks: TP= temporo-parietal. SAL-VAN = salience/ventral attention. LB = limbic. DMN = default mode. DAN = dorsal attention. SM-A =somatomotor-A. SM-B =somatomotor-B. VIS = visual.

### MDD-relevant gene expression profile

To test whether the neural markers of genetic risk for AD are linked to greater expression of MDD-relevant genes in adolescence, consistent with MDD as a precursor to AD proposal, we used the expression quantitative trait locus (eQTL) analysis data from the MDD GWAS conducted by [31]. The MDD-relevant candidate risk loci had been mapped onto the corresponding genes by the original authors using the SNP2GENE tool in FUMA and made available via the Public Results tab (https://fuma.ctglab.nl/browse). Based on their eQTL analysis output, we identified 156 MDD-linked genes reliably expressed in our data. Of these, the risk allele(s) reduced gene expression for 76 of them (MDD_low), but increased gene expression for the remaining 80 (MDD_high).

To characterise the transcriptomic signature of resilience and AD/MDD genetic risk, we focused on genes with an absolute value BSR of at least 4 (associated p-value < 10^-4^) in the brain-gene PLS analyses described above. Based on the brain-gene LV correlations, MDD-relevant transcriptomic associations with resilience, accelerated biological ageing and higher MDD PRS were derived from the number of MDD_low genes with negative BSRs and MDD_high genes with positive BSRs. Complementarily, the number of the MDD_high genes with negative BSRs and MDD_low genes with positive BSRs was used to test for an association between higher no-APOE-based AD PRS (see Figure 3-a) and stronger expression of the MDD-relevant gene expression profile.

No significant association was detected between the MDD-relevant gene expression and the resilience-linked brain LVs (*p*-values of .08 and .69 based on left hemisphere and bi-hemispheric data, respectively). In contrast, we found evidence of a significant relationship between the MDD-relevant transcriptomic profile and the neurodevelopmental profile yoked to no-APOE-based genetic risk for AD (*p*-values of .037 and .010 based on left hemisphere and bi-hemispheric data, respectively; for relative MDD gene contribution, see Figure 4-c, d).

### Receptor density maps

To elucidate the overlap between our two resilience-relevant brain profiles and density maps of neurotransmitters which play a substantial role in psychopathology (GLU/GABA [E/I], DA [combined D1/D2], HT), we used the normative group-based atlas provided by [58] and conducted a series of partial correlation analyses with permutation-based significance testing featuring 100,000 spatially constrained null brain maps [59].We thus correlated each of the two brain LVs with each of the three density maps controlling for the remaining maps (e.g., correlate brain LV1 with the combined D1/D2 receptor density map, while controlling for HT, GABA and GLU receptor densities). We identified two significant correlations replicated with a second functional atlas (see Supplemental Materials). Specifically, brain LV1 demonstrated a significant association with the DA (combined D1/D2) receptor density map (*r* of .20, spin test *p* = .046), while brain LV2 showed a significant negative correlation with the E/I density map (*r* of −.40, spin test *p* = .002) (see Figure 5-a, d for the scatter plots describing these relationships and Figure 5-b, c, e for the E/I and DA receptor density maps).

**Figure 5.**
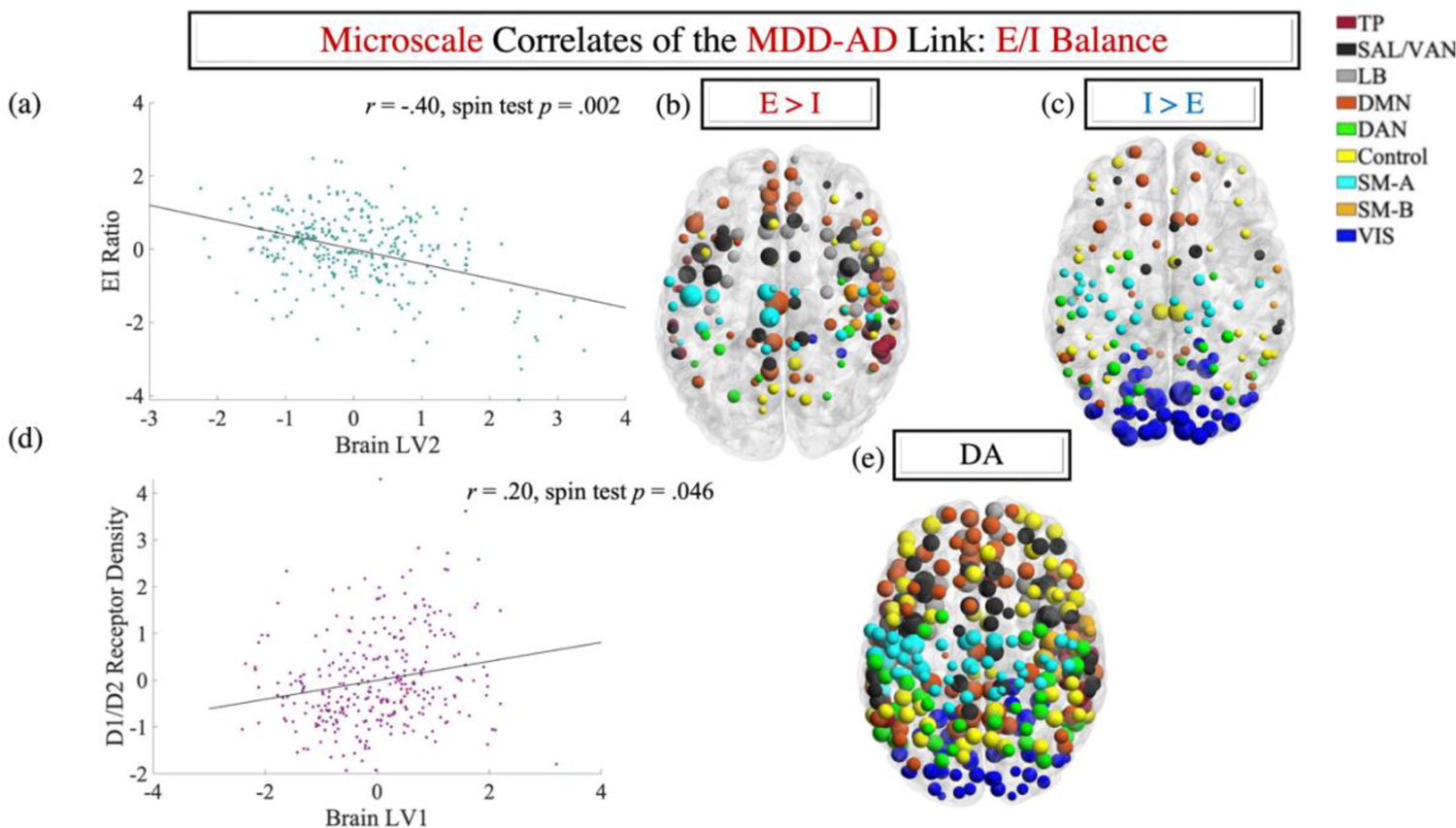
Results of the correlational analyses linking the E/I and DA receptor density map to the two brain LVs relevant to AD/MDD genetic risk, resilience and biological ageing (see Figures 2 and 3). Panels (a) and (d) contain the scatter plots describing the linear relationship between brain LV2 and the E/I (GLU/GABA) density map (a), as well as between brain LV1 and the DA density map. Panels (b) and (c) represent the areas of E > I density and I > E density, respectively, whereas panel (e) depicts the DA receptor density map. The parcels were visualized with the BrainNet Viewer (http://www.nitrc.org/projects/bnv/) (Xia et al., 2013). Parcel colours reflect Schaefer et al.’s network assignments and their size is proportional to the density of the respective receptor. E/I = excitation/inhibition. GLU = glutamate. GABA = gamma-aminobutyric acid. DA = dopamine.

### Perinatal adversity and accelerated biological ageing as moderators of the AD/MDD genetic risk-resilience relationship

Finally, we examined the role of perinatal adversity and accelerated biological ageing in modulating the impact of AD (no-APOE-based, see Figure 3-a) and/or MDD (see Figures 2-, 3-a) genetic risk on psychological resilience at the three-year follow-up. To this end, we specified the two moderated parallel mediator models conceptually represented in Figure 1. The LV2 scores in the “Follow-up Incentive/Inhibition” and “Baseline/Follow-up Incentive” conditions were robustly correlated (*r* =.43, *p* < .001). Consequently, in the absence of condition-specific predictions, the LV2 scores corresponding to the aforementioned conditions were standardised across participants and then averaged. The inhibition-linked LV1 (see Figure 2-a) and the averaged LV2 score were introduced as mediators. For the purpose of these analyses, the biological ageing variable was binned into “Low” (lowest 50% developing participants) and “High” (fastest 50% developing participants).

### Accelerated biological ageing strengthens the link between genetic risk for MDD and neurodevelopmental patterns predictive of subsequent resilience

Of the two partial moderated mediation models tested, only one emerged as significant, with an index of .014, SE = .008, 95% CI [.002; .032]. Specifically, we found that brain LV1 scores were predicted by a robust MDD PRS x biological ageing interaction, *b* = .161, SE = .064, *t*(974) = 2.510, *p* = .012 (see Figure 6-b), such that the link between the brain LV1 and MDD PRS was significant among the fastest developing participants, effect of .168, SE = .046, 95% CI [.078; .257]. Accordingly, follow-up analyses confirmed that a significant mediation of MDD PRS effects on resilience via the brain LV1 was observed only for participants showing faster biological ageing, effect of .015, SE = .007, 95% CI [.003; .030].

**Figure 6.**
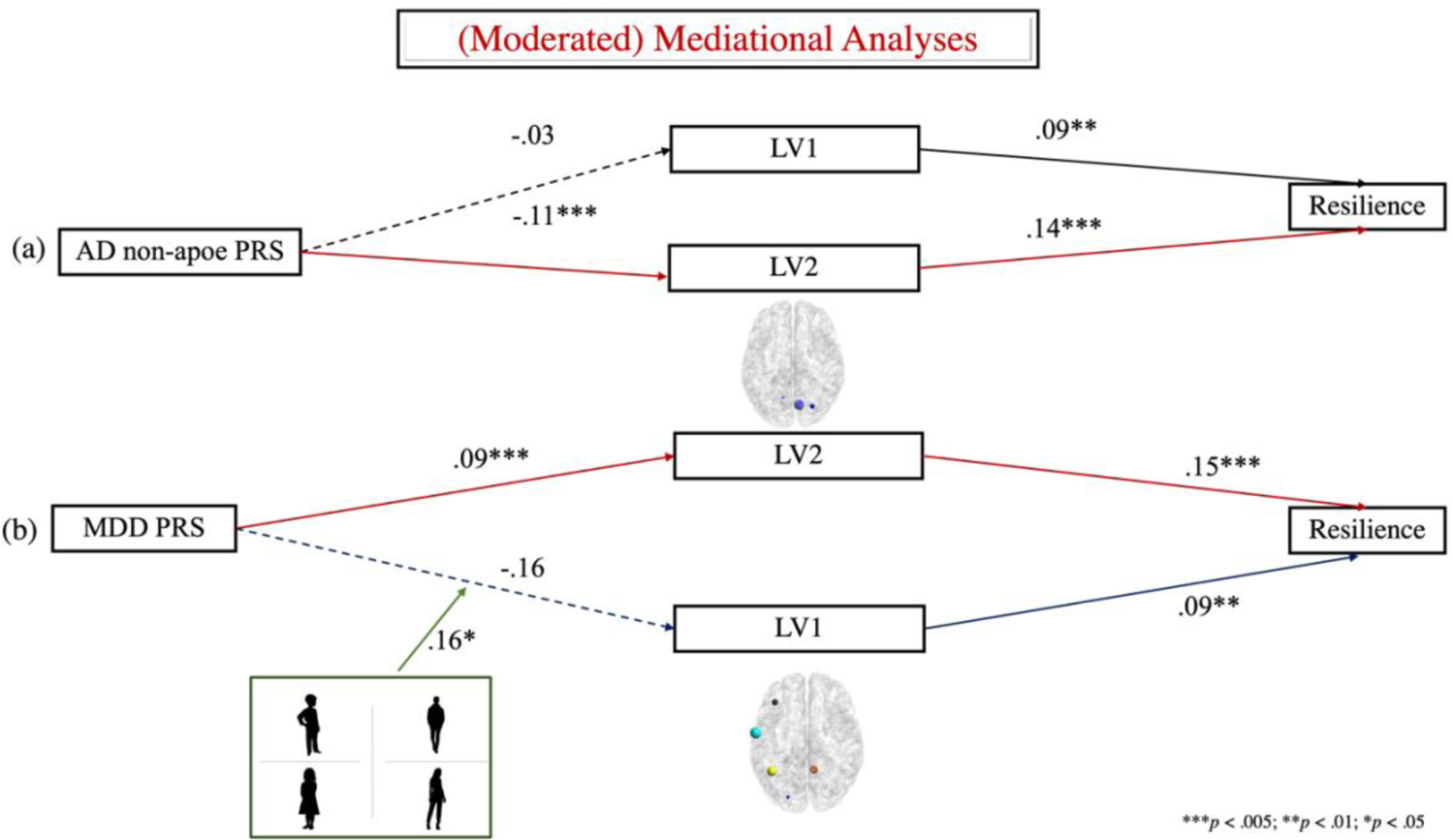
(Moderated) mediational models linking genetic vulnerability to AD (panel a) and MDD (panel b), respectively to psychological resilience as assessed at the three-year follow-up. In both panels, the coefficients associated with the red paths had been derived from simple mediational analyses (as described in the text). The coefficients associated with the blue paths are based on the moderated mediation analyses. A dashed line indicates a statistically non-significant path (*p* > .05).

### Simple mediation models

Two follow-up parallel mediator models revealed that brain LV2 partially explained the indirect effect (IE) of no-APOE-based AD vulnerability on reduced resilience, IE = −.016, SE = .006, 95% CI [-.029; −.005] (see Figure 6-a), as well as the association between the MDD PRS and greater resilience, IE = .014, SE = .006, 95% CI [.004; .027] (see Figure 6-b).

## Discussion

Applying a multidimensional approach to longitudinal data from the ABCD study, we provide novel evidence of two neurodevelopmental profiles predictive of positive adjustment to environmental challenges as a function of genetic liability to AD/MDD. One profile reflected the brain mechanisms recruited by resilient adolescents who are at risk for MDD. Reinforcing the contribution of DA to both cognitive control and resilience (42, 47, 60), this profile captured the longitudinal fine-tuning of the inhibitory control architecture, spanning areas of greater DA receptor density in functional systems that are trans-diagnostically involved in psychopathology (SAL, SM, DMN, Control, 36, 61, 62). Of note, the protective effects of this brain profile were strongest among the fastest maturing adolescents (i.e., those showing higher pubertal hormone levels), in line with prior demonstrations of DA-mediated gonadal hormone effects on behaviour, including liability to MDD (63–65). More broadly, this profile speaks to the value of longitudinal investigations of DA-gonadal hormone (particularly estradiol) dynamics in uncovering fluctuating substrates of resilience as a function of adversity exposure and differential gene expression patterns, including those observed across the various menstrual cycle stages (63, 66, 67).

A second neurodevelopmental profile differentiated resilient youth at genetic risk for MDD from psychologically vulnerable adolescents genetically predisposed towards AD. Longitudinal fine-tuning and cross-context differentiation of the neural architecture underpinning incentive processing emerged as a key resilience-promoting mechanism among the MDD-vulnerable youth. Conversely, network rigidity related to poorer resilience among AD-prone adolescents. Anchored in VIS areas of greater GABA, relative to GLU, receptor density, this profile reaffirmed the importance of externally oriented processing systems to MDD pathology (e.g., [68]), as well as the role of E/I balance in supporting functional network development (69). It also underscored the importance of conducting more targeted, neurotransmitter system-specific investigations into the relationships among AD/MDD risk, resilience and incentive processing. For instance, distinguishable GLU neuron populations in the ventral tegmental area (VTA) have been implicated in coding rewards and losses (70), while GABA and GLU neurons in the basal ganglia have been shown to code for rewards and threats, respectively (71, 72). Extending these findings, it would be worth establishing whether the observed E/I link to resilience in MDD-vs AD-predisposed individuals manifests through distinguishable pathways (e.g., threat/loss-vs reward-linked). Such a line of inquiry would be well-served by studies targeting neurotransmitter-specific substrates of whole-brain functional network interactions, while accounting for age-related modulation (73), including effective connectivity patterns between subcortical and cortical regions. This work would further benefit from experimental manipulations targeting the variety of mental processes (e.g., positive emotion upregulation vs negative emotion downregulation, 74) and cells-to-networks neurobiological mechanisms underlying resilience as a function of stressor and genetic vulnerability.

Finally, we also found evidence compatible with the putative causal role of MDD in later AD onset (10, 13). Specifically, our results raised the possibility that lower resilience among adolescents at risk for AD may stem (partly) from poorer fine-tuning of the incentive processing architecture, linked to stronger expression of MDD-relevant risk genes. As our neurodevelopmental findings centered around VIS areas, implicated in episodic memory (re)construction processes (75, 76), they are well-aligned with recent proposals on the role of episodic memory in partially explaining the link between MDD and AD genetic risk (13).

Broadly, our present findings call for more fine-grained investigations of candidate neurobiological mechanisms underpinning the link between MDD and AD. Inflammatory processes linked to hypothalamic-pituitary-adrenal (HPA) axis dysregulation (10) would warrant special attention, particularly zooming in on alterations observed in astrocytes, microglia and mitochondria, given their susceptibility to stress, E/I modulation, as well as role in neurodegeneration and psychological resilience (77–82).

### Limitations and Future Directions

Our present research opens several new avenues of inquiry. First, there is suggestive evidence that, compared to traditional laboratory tasks (such as those used in the ABCD study), naturalistic paradigms may yield neurocognitive patterns that better resemble those likely to be evoked in real-world settings (83). Consequently, replication and extension of our findings in studies featuring a movie watching paradigm, for instance, could yield further insights into the results herein documented. Second, patterns of functional network interactions underpinning optimal adjustment have been shown to vary by domain (i.e., cognition, personality, mental health) (84). As such, key insights could be gained by further probing our present findings on resilience through investigations on the neurodevelopmental mechanisms underpinning AD/MDD genetic risk and adaptive profile of cognition and/or personality.

Third, stronger genetic effects are reportedly observed on structural (relative to functional) brain indices, while structure-function coupling in connectomic features is also under substantial genetic modulation (85). Consequently, investigations of synchronised structure-function development would augment the findings herein reported, particularly given the robust predictive power of both white and gray matter indices for AD and MDD, as well as psychological resilience (86–88).

Fourth, while we provided suggestive evidence on the chemoarchitectural correlates of resilience, further cross-species investigations probing specific receptor types (e.g., D1 vs D2, 89), as well as interactions among multiple neurotransmitter systems (cf. 90) within an experimental paradigm would provide a finer grained dynamic representation of the multiscale interactions underpinning resilience. Fifth, the reliable sex differences observed in stress responses (91, 92), as well as in the genetic architecture underlying complex traits (93), highlight the need for cross-species comparative research to characterise sex-specific neurogenetic substrates of resilience, including compensatory mechanisms linked to AD/MDD vulnerability.

Finally, there is a strong need for further investigation of the mechanisms underlying the resilience-promoting effects of lifestyle choices (e.g., aerobic exercise, 94) and social relationships, including parental and peer presence in earlier life (95, 96). In particular, cross-species studies of neuroplasticity-mediated increases in resilience (97) may play a key role in designing earlier life interventions to decelerate or annihilate progression towards dementia in older adulthood.

## Conclusions

Distinguishable neurodevelopmental profiles, reflecting fine-tuning of the inhibitory control (accelerated biological ageing/DA-linked) vs incentive processing (E/I-related) architecture, predicted resilience among MDD-vulnerable adolescents. Promising support emerged for the proposed role of MDD as an early life precursor to AD, as poorer resilience among AD-vulnerable youth was associated with an MDD-relevant transcriptomic signature.

## Methods

### Participants

The sample included biological parents and offspring who participated in the ongoing Adolescent Brain Cognitive Development (ABCD) study. The analyses were based on data from Caucasian participants only because most polygenic risk markers to date have been based on this population and there is some evidence that genetic architecture and risk loci may show some racial differences (98, 99).

The present research uses baseline, two-, and three-year follow-up data downloaded in November 2021 as part of the ABCD Study Curated Annual Release 4.0 (https://data-archive.nimh.nih.gov/abcd). Following the recommendations of the ABCD study team (as detailed in the “abcd_imgincl01” file included in the data release), 980 Caucasian participants (470 female), the majority of whom were predominantly right-handed (N = 814), were selected on the basis of being biologically unrelated and having contributed high-quality data on all measures of interest. At baseline, participants were aged 9-10 years (M = 119.97 months, SD = 7.40 months). Follow-up sessions were similarly spaced across participants (baseline to two-year follow-up: M = 23.93 months, SD = 1.49 months; two- to three-year follow-up: M = 11.20 months, SD = 1.95 months).

### Psychological Resilience

Participants’ psychological resilience at the three-year follow-up was quantified by regressing out lifetime history of adverse experiences from the general psychopathology risk index (100, 101) multiplied by −1. Positive and negative residuals indicate greater and poorer resilience, respectively, relative to the sample mean (i.e., better-vs. worse-than-expected psychological functioning given experienced adversity).

### Perinatal Adversity

An index of perinatal adversity was extracted through principal components analysis from caregiver responses on the Developmental History Questionnaire (100), which was completed at baseline. This summary score, available in the ABCD 4.0 Data Release, reflects maternal prenatal care, maternal substance use during pregnancy, prenatal maternal health conditions, prematurity, birth complications and developmental milestones. The scores released by the ABCD team were multiplied by (−1), so that higher positive values would indicate greater perinatal adversity. For the purpose of our analyses, these transformed scores were subsequently binned into “Low” (lowest 50% values) and “High” (highest 50% values).

### Biological Ageing: Pubertal Timing

Parent ratings of pubertal development were selected as a measure of biological ageing due to their significant correlation with other indices of pubertal maturation. To create a more stable index of biological ageing, we averaged parent-rated pubertal development at baseline and the two-year follow-up (*r* of .65 [*p* < .001], Cronbach’s alphas of .62 and .83). While this approach to estimating biological ageing was favoured for psychometric reasons, we verified its convergence with measures of pubertal hormones, such as the dehydroepiandrosterone (DHEA), estradiol and testosterone among those participants who had both types of measures. An index of accelerated pubertal development was computed by regressing from the averaged pubertal development score the youth’s chronological age (see section on residualisation below for further details), such that a positive residual score indicated accelerated biological ageing.

### Functional Brain Architecture

Neurodevelopmental profiles relevant to inhibitory control and reward processing, respectively, were estimated during the in-scanner performance of a Stop Signal (SST) and monetary incentive delay (MID) task, respectively (26).

### MRI data acquisition and preprocessing

Acquisition parameters and preprocessing steps for the ABCD study data are described in [102]. Our analyses used minimally preprocessed fMRI data available as part of the ABCD Study Curated Annual Release 4.0. The minimal preprocessing pipeline involved correction for head motion spatial and gradient distortions, bias field removal, and co-registration of the functional images to the participant’s T1– weighted structural image. We further applied the following steps: (1) elimination of initial volumes (8 volumes [Siemens, Philips], 5 volumes [GE DV25], 16 volumes [GE DV26] to allow the MR signal to reach steady state equilibrium, (2) linear regression-based removal of quadratic trends and 24 motion terms (i.e., the six motion parameters, their first derivatives, and squares, cf. 151) from each voxel’s time course.

### Parcel definition

Our main analyses were based on the Schaefer 300 parcel-functional atlas (51), downloaded from https://github.com/ThomasYeoLab/CBIG. The atlas version we used encompasses 17 functional networks, spanning core systems, such as the DMN (A/B/C), Control (A/B/C), Salience/Ventral Attention (A/B), Dorsal Attention (DAN A/B), Somatomotor (SM A/B), Visual (VIS Central/Peripheral), Limbic (LB A/B), and Temporo-parietal (TP). The Schaefer atlas was available in the Montreal Neurological Institute (MNI) standard space. To align it with the participants’ native space for each of the four tasks (SST at Time 1/Time 2, MID at Time 1/Time 2), the following steps were implemented in FSL: (1) the middle image in each task run was converted to the MNI space (via the MNI-152 brain template available in FSL) and the inverse transformation (MNI-to-participant native space) was estimated; (2) the inverse transformation was used to align the Schaefer atlas to each participant’s functional images, separately for each task run (SST at Time 1/Time 2, MID at Time 1/Time 2).

### Parcel-to-parcel correlations in timeseries

Pairwise Pearson’s correlations between all the Schaefer parcels were computed separately for the SST and MID in Matlab and expressed as Fisher’s z-transformed scores. Because the two tasks are very similar in duration, we used all available data from each. Similar to prior studies using multilayer community detection algorithms (e.g., 53), only the positive Fisher’s z-scores were entered in the network-level analyses detailed below, while negative z-scores were set to zero.

### Network-level analyses

All the network-level metrics were computed using the Network Community Toolbox (52), as described below. Patterns of parcel-based functional reorganisation, longitudinally within each task and between the two tasks at the same time point were characterised with a multilayer generalised Louvain-like community detection algorithm implemented in the NCT. In line with extant practices (53), the spatial resolution parameter was set to the default value of 1. Taking our cue from other investigations of heterogenous mental states (53), we set to 0.5 the cross-layer (MID-SST at Time 1; MID-SST at Time 2; Time 1-to-Time 2 MID; Time 1-to-Time 2 SST) connection strength parameter. To account for the near degeneracy of the modularity landscape, the multilayer community detection algorithm was initiated 100 times and all the functional network interactions indices detailed below were averaged across all iterations.

### Functional network development

Longitudinal (baseline-to-2-year follow-up) within-task and cross-task (SST/MID) contemporaneous differences in functional brain architecture were estimated across all the modularity optimisation iterations. Specifically, using the “flexibility” function in NCT, we quantified the number of times each parcel in the Schaefer atlas changed communities between the two tasks at each wave and, within each task, from baseline to follow-up.

### Polygenic Risk Scores (PRS)

MDD and AD PRSs were each computed as the weighted sum of risk alleles based on the summary statistics of two large GWASs focused on each disorder (30, 31, for AD and MDD, respectively). We focused on these two GWASs for the following reasons: (1) public availability of eQTL analysis output; (2) match to our Caucasian-only sample with [31] in particular, being the largest MDD GWAS to date of this population; (3) use of clinical diagnosis-based patient cases and, for [30] confirmed clinical cases of late-onset sporadic AD, which would ease the interpretation of the derived PRSs and associated findings.

For AD, we computed a separate APOE region (chromosome 19:44.4-46.5 Mb)-vs no-APOE region PRS because the two PRSs forecast distinguishable trajectories of neurocognitive impairments and differential susceptibility to environmental factors (16). For MDD, we used the the top 10k most informative variants, based on approximately 76k patients and 230k controls, which had been made publicly available by [31]. These variants had been obtained by clumping the corrsponding GWAS statistics with the following parameters p1 = p2 = 1, window size < 500kb, and r^2^ > 0.1.

Prior to PRS computation, the following preprocessing steps were implemented: (1) genes with a minor allele frequency (MAF) < .05, insertion/deletion and ambiguous single nucleotide polymorphisms (SNPs) (i.e., A/T and G/C pairs) were excluded; (2) highly correlated SNPs (r^2^ > .10) within a 500 kb window were eliminated. The SNPs which survived the preprocessing and had an associated GWAS level *p* ≤ 5×10^-8^ contributed to the computation of the disorder-specific PRSs (MDD PRS: N = 8 SNPs; no-APOE AD PRS: N = 10 SNPs; APOE AD PRS = 14 SNPs). In our main analyses we opted to use a stringent *p*-value (i.e., GWAS-level significant at *p* < .05) in order to identify the variants contributing most robustly to predicting genetic risk for AD/MDD. However, in supplemental analyses, we confirmed that the identified brain patterns linked to AD/MDD risk and resilience also emerged when using more lenient *p*-thresholds (see Figures S3-6).

### Gene expression data processing and analysis

#### Microarray gene expression

Micro-array gene expression data were obtained from six postmortem brains (1 female, ages 24.0–57.0, 42.50 +/- 13.38) provided by the Allen Institute for Brain Science (https://www.brain-map.org/). Because all six brains had left hemisphere data, but only two brains contained data from the right hemisphere, our gene-brain analyses focused on the left hemisphere parcels. However, consistent with observations that gene expression patterns are largely symmetric across the two hemispheres, we provide evidence that our results are replicated when using gene expression patterns mirrored across the two hemispheres.

The gene expression data was processed with abagen (https://github.com/netneurolab/abagen), following the steps outlined in [57]. The resulting gene expression matrices, used in all our analyses, were in the format 297 (bilateral)/147 (left-hemisphere) (parcels) x 15,632 (genes). A list of parcels lacking reliable gene expression is included in the Supplemental Materials (Table S1).

#### MDD overlap tests

To test whether the neural markers of genetic risk for AD are linked to greater expression of MDD-relevant genes in adolescence, consistent with MDD as a precursor to AD proposal, we used the eQTL analysis data from the MDD GWAS conducted by [31]. The MDD-relevant candidate risk loci had been mapped onto the corresponding genes by the original authors using the SNP2GENE tool in FUMA and made available via the Public Results tab (https://fuma.ctglab.nl/browse). Based on their eQTL analysis output, we identified 156 MDD-linked genes reliably expressed in our data. Of these, the risk allele(s) reduced gene expression for 76 of them (MDD_low), but increased gene expression for the remaining 80 (MDD_high).

To characterise the transcriptomic signature of resilience and AD/MDD genetic risk, we focused on genes with an absolute value BSR of at least 4 (associated p-value < 10^-4^) in the brain-gene PLS analyses. Comparisons were conducted separately for genes with positive versus negative loadings on the gene LV. As an example, for the positive BSR genes, the procedure was as follows: (1) we obtained separate counts of the number of MDD_low and MDD_high genes, respectively, with a BSR of at least 4 on our gene LV (MDD_low_pos and MDD_high_pos, respectively); (2) we counted the number of genes with a BSR of at least 4 on our gene LV (Orig_pos); (3) from each of the corresponding gene LVs in the null distribution (each of which had been aligned with the original gene LVs via a Procrustes transform), we selected a number of genes equal to Orig_pos (Null_pos); (4) we counted the number of null samples (out of the total of 100,000) in which the number of either MDD_low_pos or MDD_high_pos in Null_Pos exceeded the one observed in Orig_pos. The same procedure was followed for the negative BSR genes. Estimation of whether the identified transcriptomic signatures implied greater risk for MDD varied depending on whether the brain and gene were positively versus negatively correlated. For example, if the gene and brain LVs were positively correlated, MDD risk would be estimated as a conjunction of MDD_low genes with negative BSRs and MDD_high genes with positive BSRs observed in the original data relative to the null distribution.

#### Control analyses: AD-relevant transcriptomic signature

The very low number (=4) of AD risk genes in the thresholded (BSR > 4) left-hemispheric gene data prevented us from running any control analyses relevant to the transcriptomic signature of AD. For the analyses using bi-hemispheric data, we found no evidence of a statistically significant link between the AD-relevant transcriptomic profile and either of the two identified brain LVs (*p*s > .07).

### Receptor Density Maps: DA, GABA, GLU, HT

To estimate receptor density maps for our neurotransmitters of interest (DA, GABA, GLU, HT), we used the normative atlas put together by [58], which is based on positron emission tomography (PET) data from over 1200 healthy individuals. Group-averaged pre-processed PET images corresponding to each tracer map for DA, GABA, GLU, HT were downloaded from https://github.com/netneurolab/hansen_receptors/tree/main/data/PET_nifti_images. Each tracer map was parcellated in the MNI-152 space based on the Schaefer atlas using the “Parcellater” function from neuromaps https://github.com/netneurolab/neuromaps/tree/main/neuromaps. Following the strategy from [58], an adapted version of the “make_receptor_matrix.py” script (https://github.com/netneurolab/hansen_receptors/tree/main/code) was employed to estimate the weighted average of the receptor density maps corresponding to each neurotransmitter of interest. This was done separately for each tracer based on the number of participants in the respective study (see Table 1 in [58]).

Each receptor density measure was standardised across all the parcels in the Schaefer atlas. The multiple indices of DA and HT receptor density were moderately to strongly correlated (DA [D1/D2]: *r* of .52; HT: *r*s from .28 to .57). Consequently, a global index of DA (combined D1/D2) and HT, respectively, receptor density was computed by averaging the standardised scores of the corresponding indices. A parcel-specific E/I measure was created by subtracting the GABA receptor density standardised score from the GLU receptor density standardised score. Brain regions with higher values on this index show relatively greater GLU (compared to GABA) receptor density.

### Control Variables

To characterise the underpinnings of resilience in relation to the total number of acute stressful experiences, we controlled for chronic exposure to material deprivation, family conflict and neighbourhood crime, all of which had been shown to impact inhibitory control, incentive processing and susceptibility to psychopathology.

### Residualisation

The non-imaging variables were residualised for the following confounders:

1. chronological age in order to estimate accelerated/decelerated development;
2. biological sex (coded as “1” for females, “0” for males);
3. handedness (coded as “0” for right-handedness and “1” for non-right-handedness);
4. serious medical problems, which was based on the ABCD Parent Medical History Questionnaire and computed as an average of unplanned hospital visits in the prior year for chronic health conditions, head trauma, loss of consciousness and/or convulsions/trauma;
5. scanner site (21 dummy variables to account for scanner-related differences, as well as broad differences in family education and socio-economic status across sites);
6. material deprivation, family conflict and neighbourhood crime;
7. average modality-specific motion per participant (151);
8. difference (in months) between the baseline and two-year follow-up sessions (only for the non-imaging variables involved in longitudinal comparisons).

Because the difficulty of the MID and SST trials was dynamically adjusted to maintain a set accuracy level (26), we saw no reason to control for behavioural performance.

### Data reduction: Multivariate normality

Because the multiple linear regression analyses used for confounder residualisation are sensitive to violations of multivariate normality, a square-root transformation was applied to the Total Problem scores (which contributes to the psychological resilience index) and the APOE-based AD PRS scores prior to any analyses. Subsequently, an examination of the PRSs, resilience and accelerated biological aging residuals based on the normal Q-Q plots and the Kolmogorov-Smirnov test (with the Lilliefors significance correction) confirmed the multivariate normality of all non-imaging variables under scrutiny.

### fMRI and PRS Data Analysis

#### Partial least squares analysis (PLS)

Partial least squares correlation (i.e., PLS), a multivariate data-driven manner technique which can identify relationships between neural patterns (latent variables or LVs) and individual differences variables (behavioural PLS), was used to probe the transcriptomic and functional brain profiles linked to AD/MDD-related genetic vulnerability, psychological resilience and accelerated biological ageing.

Two sets of behavioural PLS analyses were conducted. The sole analysis in the first set featured resilience, biological ageing, as well as the MDD and AD (APOE-vs non-APOE-based) PRSs in the “behavioural” matrix. The brain matrix contained the flexibility indices estimated in NCT and modelled as four separate conditions reflecting functional brain reorganisation, longitudinally within each task (incentive vs inhibitory control), as well as between the two tasks at baseline and the two-year follow-up, respectively. The second set of PLS analyses sought to characterise the transcriptomic signature of resilience, biological ageing and AD/MDD genetic vulnerability. Specifically, it estimated the correlation between the brain LVs identified in behavioural PLS analysis 1 and the parcel x gene expression level matrix derived with the abagen toolbox from the comprehensive transcriptomic maps provided by the Allen Brain Institute.

#### Significance and reliability testing

In all the reported PLS analyses, the significance of each LV was determined using a permutation test (5000 permutations for the brain-[behavior] analyses and 100,000 permutations for all the analyses involving gene expression data). In the gene-brain PLS analysis, to account for correlated gene expression patterns based on anatomical proximity we used Vasa’s “rotate_parcellation” function in Matlab (https://github.com/frantisekvasa/rotate_parcellation/commit/bb8b0ef10980f162793cc180cef371e83655c505) in order to generate 100,000 spatially constrained permutations of the Schaefer brain LV, as identified in behavioral PLS analysis 1. These spatially constrained permuted brain LVs were used to assess the significance of the extracted gene LVs.

The reliability of each parcel’s contribution to a particular LV was tested by submitting all weights to a bootstrap estimation (1000 bootstraps for the brain-[behaviour] analyses and 100000 bootstraps for all the analyses involving gene expression data) of the standard errors (SEs) A bootstrap ratio (BSR) (weight/SE) of at least 3 in absolute value (approximate associated p-value < .005) was used as a threshold for identifying those parcels that made a significant contribution to the identified LVs. For the gene PLS analyses, we focused on approximately the top 10% of absolute value BSRs (i.e., ∼4, associated *p*-value < 10^-4^).

### Mediation analyses

To test whether perinatal adversity and/or accelerated biological ageing modulate the neurodevelopmental correlates of resilience as a function of genetic risk for AD and MDD, we conducted two moderated mediation analyses using Hayes’ PROCESS 3.5 macro for SPSS. These analyses were based on the PLS results and, as such, for AD, they only involved the no-APOE-based PRS. Two simple mediation analyses involving the same predictors, mediators and outcome were also performed for exploratory purposes.

## Data Availability

The raw data are available at https://nda.nih.gov/abcd upon completion of the relevant data use agreements. The ABCD data repository grows and changes over time. The ABCD data used in this report came from Adolescent Brain Cognitive Development Study (ABCD) - Annual Release 4.0 #1299. DOIs can be found at http://dx.doi.org/10.15154/1523041

## Acknowledgments

Data used in the preparation of this article were obtained from the Adolescent Brain Cognitive Development (ABCD) Study (https://abcdstudy.org), held in the NIMH Data Archive (NDA). This is a multisite, longitudinal study designed to recruit more than 10,000 children age 9-10 and follow them over 10 years into early adulthood. The ABCD Study is supported by the National Institutes of Health and additional federal partners under award numbers U01DA041048, U01DA050989, U01DA051016, U01DA041022, U01DA051018, U01DA051037, U01DA050987, U01DA041174, U01DA041106, U01DA041117, U01DA041028, U01DA041134, U01DA050988, U01DA051039, U01DA041156, U01DA041025, U01DA041120, U01DA051038, U01DA041148, U01DA041093, U01DA041089, U24DA041123, U24DA041147. A full list of supporters is available at https://abcdstudy.org/federal-partners.html. A listing of participating sites and a complete listing of the study investigators can be found at https://abcdstudy.org/consortium_members/. ABCD consortium investigators designed and implemented the study and/or provided data but did not participate in analysis or writing of this report. This manuscript reflects the views of the authors and may not reflect the opinions or views of the NIH or ABCD consortium investigators. The authors would like to thank Valentina Escott-Price for advice on the polygenic risk analyses.

## Materials & Correspondence

Correspondence and material requests should be addressed to R.P.

## Code availability

We used already existing code, as specified in the main text with links for free download.

