## Supplementary material for "Psychological Resilience in Adolescence as a function of Genetic Risk for Major Depressive Disorder and Alzheimer’s Disease": SI

### Supplemental Materials

#### Methods

##### Participants

The sample included biological parents and offspring who participated in the ongoing Adolescent Brain Cognitive Development (ABCD) study (for a detailed sample description, see [1]). The analyses were based on data from Caucasian participants only because most polygenic risk markers to date have been based on this population and there is some evidence that genetic architecture and risk loci may show some racial differences (2-3).

The present research uses baseline, two-, and three-year follow-up data downloaded in November 2021 as part of the ABCD Study Curated Annual Release 4.0 (https://data-archive.nimh.nih.gov/abcd). Following the recommendations of the ABCD study team (as detailed in the “abcd_imgincl01” file included in the data release), 980 Caucasian participants (470 female), the majority of whom were predominantly right-handed (N = 814), were selected on the basis of being biologically unrelated and having contributed high-quality data on all measures of interest. At baseline, participants were aged 9-10 years (M = 119.97 months, SD = 7.40 months). Follow-up sessions were similarly spaced across participants (baseline to two-year follow-up: M = 23.93 months, SD = 1.49 months; two- to three-year follow-up: M = 11.20 months, SD = 1.95 months).

##### Psychological Resilience

Participants’ psychological resilience at the three-year follow-up was quantified by regressing out lifetime history of adverse experiences from the general psychopathology risk index (as described below) multiplied by -1. Positive and negative residuals indicate greater and poorer resilience, respectively, relative to the sample mean (i.e., better- vs. worse-than-expected psychological functioning given experienced adversity).

**Psychopathology Risk: Child Behaviour Checklist (CBCL).** Global psychopathology risk was quantified with the Total Problems score derived from the parent-report version of the CBCL (4), which is available in the ABCD 4.0 Data Release. This 112-item instrument uses a 3-point Likert rating scale (0 = not true, 1 = somewhat or sometimes true, 2= very true or often true) to gauge participants' psychological functioning in the prior six months. The Total Problems score was computed as a sum of the response values on all items (no missing values were recorded in our sample). In line with extant guidelines (5,6), raw scores (rather than t-scores) were used given their greater precision particularly at the extremes of the scale. In the present study, this strategy was deemed particularly appropriate, since chronological age and biological sex were controlled for in all analyses.

**Lifetime History of Adverse Life Events.** At the 3-year follow-up, lifetime exposure to negative events was estimated with an adapted version of the 25-item Adverse Life Events scale, which further incorporated questions about the caregiver being hospitalised, youth being put in foster care, youth seeing someone been beaten up or shot at in school/neighbourhood and having a lockdown in school due to concerns over violence (7). The scale used a Yes/No response format to gauge event occurrence, accompanied by two 4-point scales assessing event valence and impact, respectively. Due to the relatively low completion rate of the event rating scales, our analyses focused on the lifetime event occurrence for which there were no missing data. Youth, rather than parent, reports were used for two reasons. First, we sought to elucidate the correlates of resilience in response to consciously experienced adversity. Second, we intended to minimise measurement bias in estimating resilience by having a different informant rate adversity exposure (i.e., child) versus psychopathology risk (i.e., parent).

**Perinatal Adversity**

An index of perinatal adversity was extracted through principal components analysis from caregiver responses on the Developmental History Questionnaire (8), which was completed at baseline. This summary score, available in the ABCD 4.0 Data Release, reflects maternal prenatal care, maternal substance use during pregnancy, prenatal maternal health conditions, prematurity, birth complications and developmental milestones. The perinatal adversity factor correlated most strongly with birth weight and prematurity, two core stressors predictive of subsequent brain-wide alterations in normative development (9-13).

The scores released by the ABCD team were multiplied by (-1), so that higher positive values would indicate greater perinatal adversity. For the purpose of our analyses, these transformed scores were subsequently binned into “Low” (lowest 50% values) and ”High” (highest 50% values).

**Biological Ageing: Pubertal Timing**

The 5-item Pubertal Development Scale (PDS) was selected as a measure of biological ageing due to its significant correlation with other indices of pubertal maturation, including physician ratings (14). This questionnaire uses a 4-point Likert type response format, ranging from 1 (no development) to 4 (development already completed) to assess development along three gender-general (i.e., growth spurt, changes in skin, hair growth) and two gender-specific (i.e., facial hair growth, voice change [boys]; breast development, menarche [girls]) dimensions. For reasons of data completeness and scale reliability at baseline, our analyses focused on parent ratings of pubertal development. To create a more stable index of biological ageing, we averaged parent-rated PDS scores at baseline and the two-year follow-up ($r$ of .65 [$p < .001$], Cronbach’s alphas of .62 and .83). While this approach to estimating biological ageing was favoured for psychometric reasons (as outlined above) and due to the ease of drawing connections with the prior literature (cf. 15), we verified its convergence with measures of pubertal hormones, such as the dehydroepiandrosterone (DHEA), estradiol and testosterone (cf. 16) among those participants who had both types of measures. To this end, we standardised and then averaged the baseline and two-year mean measurements for each hormone. Partial correlational analyses, controlling for potential confounds at each wave, such as caffeine consumption and activity levels within the 12 hours preceding measurement, time since midnight, collection duration and time to freeze (cf. 16), revealed the expected positive association of parent-rated PDS scores with levels of DHEA, $r(933) = .34$, $p < .001$, and testosterone, $r(933) = .29$, $p < .001$ across both genders, as well as levels of estradiol among females, $r(440) = .15$, $p = .002$.

An index of accelerated pubertal development was computed by regressing from the averaged PDS score the youth's chronological age (see section on residualisation below for further details), such that a positive residual score indicated accelerated biological ageing (cf. 15).

**Functional Brain Architecture**

**Inhibitory control.** An in-scanner Stop Signal Task (SST) measured the ability to inhibit an ongoing speeded motor response to a "Go" signal. In ABCD, the task comprises two runs of 180 trials each: 150 "Go" trials, 15 "Stop" trials expected to be unsuccessful and 15 "Stop" trials expected to be successful. To maintain the breakdown of the successful/unsuccessful "Stop" trials, a tracking algorithm was implemented to alter the interval between the presentation of the 'Go stimulus' and the onset of the 'Stop' signal based on the participant's performance. Each run was restricted to begin with a 'Go' trial and stop trials were separated by a minimum of one 'Go' trial (for further details on this task, including criticisms pertaining mostly to behavioural data analyses, see 17).

**Incentive Processing.** A monetary incentive delay task gauged anticipatory and consummatory reactions to rewards and losses, as well as drive to engage in speeded responses for monetary gains or avoidance of losses. Each of the two task runs contains 50 contiguous trials consisting of a monetary incentive cue presented for 2000 ms (10 of each type: Win $.20, Win $5, Lose $.20, Lose $5, $0-no money), a variable (1500-4000 ms) anticipation period, a response-to-target interval (150-500ms), followed by feedback on the outcome of the trial (2000 ms – target duration). In total, there are 40 reward, 40 loss and 20 no-money trials. Task parameters are adjusted for each individual participant in order to maintain an overall accuracy of 60% (17).

**MRI data acquisition.** Scanning was performed across 21 US sites, with a protocol harmonised for Siemens Prisma, Philips, and GE 3T scanners (for details, see 17,18). Scanner type was controlled for in all analyses by using site id as a covariate to account for magnet and sociodemographic differences among sites (150). T1-weighted were acquired with an MPRAGE-PMC (Prospective Motion Correction) sequence (TR=2500 (Siemens/GE)/6.31 (Phillips) ms, TE= 2.88 (Siemens)/2.9 (Phillips)/2 (GE) ms, flip angle=8°, FOV = 256 x 256 mm, 176 (Siemens)/225 (Phillips)/208 (GE) slices of 1 × 1 mm in-plane resolution, 1 mm thick). The fMRI data were acquired with a multiband EPI sequence (TR=800 ms, TE=30 ms, flip angle=52°, FOV = 216 x 216 mm, 60 slices of 2.4 × 2.4 mm in-plane resolution, 2.4 mm thick, multiband acceleration factor of 6).

**Network-level analyses.** All the network-level metrics were computed using the Network Community Toolbox (NCT, Bassett, D.S. [2017, November]. Network Community Toolbox. Retrieved from http://commdetect.weebly.com/), as described below.

To characterise patterns of parcel-based functional reorganisation between the two tasks at the same time point and, longitudinally, within the same task, we used a multilayer generalised Louvain-like community detection algorithm, first introduced by [21] and implemented in the NCT. This algorithm partitions a network with multiple layers into non-overlapping groups of nodes (i.e., functional communities) with the goal of maximising an objective modularity quality function, defined as

$$\mathbf{Q} = \frac{1}{2\mu}\sum_{ijlr}[(w_{ijl} - \gamma_l e_{ijl})\delta_{lr} + \delta_{ij}\omega_{jlr}]\delta(g_{il}, g_{jr})$$

where 2μ is the sum of all connection weights in the network across all layers, $w_{ijl}$ represents the connection strength between nodes i and j in layer l; $\gamma_l$ is a resolution parameter determining the size of the identified modules in layer l; $e_{ijl}$ is the connection strength expected by chance between nodes i and j in layer l, and defined as $e_{ijl} = \frac{s_{il}s_{jl}}{v}$ with $s_{il}$ and $s_{jl}$ being the sum of all connection weights of node i and j, respectively, in layer l, while v is the sum of all connection weights in the network in layer l; $\omega_{jlr}$ is the connection strength between node j in layer l and node j in layer r, and $g_{il}$ and $g_{jr}$ give the community assignments of node i in layer l and node j in layer r.

In the above modularity quality optimisation, there are two free parameters, the spatial resolution parameter, γ, which tunes community size within each layer, and the cross-layer connection strength parameter, ω, which determines community stability across layers. In line with extant practices (20, 22), the spatial resolution parameter was set to the default value of 1. Taking our cue from other investigations of heterogenous mental states (20), we set to 0.5 the cross-layer (MID-SST at Time 1; MID-SST at Time 2; Time 1-to-Time 2 MID; Time 1-to-Time 2 SST) connection strength parameter. To account for the near degeneracy of the modularity landscape (23), the multilayer community detection algorithm was initiated 100 times and all the functional network interactions indices detailed below were averaged across all iterations.

***Functional network development.*** Longitudinal (baseline-to-2-year follow-up) within-task and cross-task (SST/MID) contemporaneous differences in functional brain architecture were estimated across all the modularity optimisation iterations. Specifically, using the “flexibility” function in NCT, we quantified the number of times each parcel in the Schaefer atlas changed communities between the two tasks at each wave and, within each task, from baseline to follow-up.

**Polygenic Risk Scores (PRS)**

MDD and AD PRSs were each computed as the weighted sum of risk alleles based on the summary statistics of two large GWASs focused on each disorder (24, 25, for MDD and AD, respectively), which had been made available by the original authors via the “Public Results” tab on the FUMA website (https://fuma.ctglab.nl/browse, 26).

For AD, we computed a separate APOE region (chromosome 19:44.4-46.5 Mb)- vs no-APOE region PRS (cf. 27) because the two PRSs forecast distinguishable trajectories of neurocognitive impairments and differential susceptibility to environmental factors, including stress (28, 29). For MDD, we used the top 10k most informative variants, based on approximately 76k patients and 230k controls, which had been made publicly available by [24] on FUMA (including the associated eQTL analysis output). These variants had been obtained by clumping the corresponding GWAS statistics with the following parameters p1 = p2 = 1, window size < 500kb, and r2 > 0.1.

PRS computation based on the *.genotype ABCD data followed established guidelines (30, 31). Specifically, the following preprocessing steps were implemented: (1) genes with a minor allele frequency (MAF) < .05, insertion/deletion and ambiguous single nucleotide polymorphisms (SNPs) (i.e., A/T and G/C pairs) were excluded; (2) highly correlated SNPs ($r^2 > .10$) within a 500 kb window were eliminated. The PRSs used in our main analyses were based on GWAS significant SNPS (i.e., $p$-value $\leq 5x10^{-8}$ ; MDD PRS: N = 8 SNPs; no-APOE AD PRS: N = 10 SNPs; APOE AD PRS = 14 SNPs) since these are likely to make the most robust contribution to disease risk. However, in supplemental analyses, we confirmed that that identified brain patterns linked to AD/MDD risk and resilience also emerge when using more lenient *p*-thresholds (see Figures S3-6).

**Gene expression data processing and analysis**

**Microarray gene expression.** Micro-array gene expression data were obtained from six postmortem brains (1 female, ages 24.0–57.0, 42.50 +/- 13.38) provided by the Allen Institute for Brain Science (https://www.brain-map.org/). Because all six brains had left hemisphere data, but only two brains contained data from the right hemisphere, our gene-brain analyses focused on the left hemisphere parcels (cf. 32). However, consistent with observations that gene expression patterns are largely symmetric across the two hemispheres, we provide evidence that our results are replicated when using gene expression patterns mirrored across the two hemispheres.

The gene expression data was processed with abagen (https://github.com/netneurolab/abagen). Microarray probes were reannotated based on the probe-to-gene mapping information provided by [33] and filtered based on their expression intensity relative to background noise, such that probes with intensity less than the background in >=50% of samples across donors were discarded. When multiple probes indexed the expression of the same gene, we selected and used the probe with the most consistent pattern of regional variation across donors (i.e., differential stability; 34). The MNI coordinates of tissue samples were updated to those generated via non-linear registration using the Advanced Normalization Tools (ANTs; https://github.com/chrisfilo/alleninf). Samples were assigned to brain regions in the Schaefer atlas if their MNI coordinates were within 2 mm of a given parcel. All tissue samples not assigned to a brain region in the provided atlas were discarded.

Inter-subject variation was addressed by normalizing tissue sample expression values across genes using a robust sigmoid function (35):

$xnorm = 1/(1+\exp(-(x-\langle x\rangle)/\mathrm{IQR}x))$

where $\langle x\rangle$ is the median and IQR$x$ is the normalized interquartile range of the expression of a single tissue sample across genes. Normalized expression values were then rescaled to the unit interval:

$xscaled = (xnorm - \min(xnorm))/(\max(xnorm) - \min(xnorm))$

Gene expression values were then normalized across tissue samples using an identical procedure. Samples assigned to the same brain region were averaged separately for each donor and then across donors. After we eliminated the parcels without reliable gene expression (based on the abagen parameters outlined above), the resulting gene expression matrices, used in all our analyses, were in the format 297 (bilateral)/147 (left-hemisphere) (parcels) x 15,632 (genes). A list of parcels lacking reliable gene expression is included in the Supplemental Materials (Table S1).

Profiles of neural excitation/inhibition (E/I) (im)balance are reportedly foundational to the cognitive control deficits that typify both AD and MDD (38). Consequently, a parcel-specific E/I measure was created by subtracting the GABA receptor density standardised score from the GLU receptor density standardised score. Brain regions with higher values on this index show relatively greater GLU (compared to GABA) receptor density.

**Control Variables**

To characterise the underpinnings of resilience in relation to the total number of acute stressful experiences, we controlled for chronic exposure to the risk factors described below, which had been shown to impact inhibitory control, incentive processing and susceptibility to psychopathology (cf. 15, 39).

**Material deprivation.** Financial deprivation was assessed with a 7-item scale developed by [40] to assess unmet material needs in the areas of housing, food and medical care in the 12 months preceding assessment. Each item is scored as 1 or 0 (yes/no). Responses to all items were averaged with higher scores indicating experiences of greater financial hardship. To create a more stable index of financial deprivation covering the baseline to the three-year follow-up interval, we averaged the aggregate scale scores corresponding to the yearly assessments (interwave *r*s from to .43 to .55; Cronbach's alphas from .69 to .75; M = .23 [SD = .63] across all waves).

**Exposure to violence.** The exposure to violence measures described below were completed independently by the parent and the youth. One participant lacked parent ratings of family conflict and neighbourhood crime at the two-year follow-up, hence, in this case, the follow-up youth ratings were used instead.

***Family conflict***. A 9-item Family Conflict scale [41] gauged exposure to domestic violence. Each item is scored as 1 or 0 for true/false, with reverse coding of items that imply lack of conflict in the home (e.g., "We fight a lot in our family." versus "Family members rarely become openly angry."). Higher scores indicate a more conflictual family environment. Both parent (Cronbach's alphas of .66 [M = .25, SD = .21] and .70 [M = .26, SD = .22] for baseline and two-year follow-up, respectively) and youth (Cronbach's alphas of .65 [M = .21, SD = .20] and .66 [M = .18, SD = .20] for baseline and two-year follow-up, respectively) versions demonstrated acceptable reliability.

***Perceived neighbourhood crime and safety***. The 3-item Neighbourhood Safety/Crime Scale from [42] uses a five-point Likert Scale (5 = strongly agree to 1= strongly disagree) to gauge perceptions of threat related to the neighbourhood in which the respondent resides (i.e., areas within a 20-minute walk from the respondent's home). At both time points, the youth completed only a 1-item version of the scale ("My neighbourhood is safe from crime"; M = 4.22 [SD = .93] and M = 4.22 [SD = .90] for baseline and two-year follow-up, respectively), whereas the parent filled out the full 3-item version of the scale (Cronbach's alphas of .85 [M = 4.09, SD = .82] and .82 [M = 4.07, SD = .78] for baseline and two-year follow-up, respectively). Higher scores on this scale indicate greater neighbourhood safety.

**Residualisation**

To minimise bias in our multivariate brain-behaviour analyses (cf. 43), only the non-imaging variables were residualised for the following confounders:

(1) chronological age in order to estimate accelerated/decelerated development;

(1) biological sex (coded as "1" for females, "0" for males);

(2) handedness (coded as "0" for right-handedness and "1" for non-right-handedness);

(3) serious medical problems, which was based on the ABCD Parent Medical History Questionnaire and computed as an average of unplanned hospital visits in the prior year for chronic health conditions, head trauma, loss of consciousness and/or convulsions/trauma;

(4) scanner site (21 dummy variables to account for scanner-related differences, as well as broad differences in family education and socio-economic status across sites);

(5) material deprivation, family conflict and neighbourhood crime;

(6) average modality-specific motion per participant;

(7) difference (in months) between the baseline and two-year follow-up sessions (only for the non-imaging variables involved in longitudinal comparisons).

Because the difficulty of the MID and SST trials was dynamically adjusted to maintain a set accuracy level (17), we saw no reason to control for behavioural performance.

**Data reduction: Multivariate normality.** Because the multiple linear regression analyses used for confounder residualisation are sensitive to violations of multivariate normality (44), a square-root transformation was applied to the Total Problem scores (which contributes to the psychological resilience index) and the APOE-based AD PRS scores prior to any analyses. Subsequently, an examination of the PRSs, resilience and accelerated biological aging residuals based on the normal Q-Q plots and the Kolmogorov-Smirnov test (with the Lilliefors significance correction) confirmed the multivariate normality of all non-imaging variables under scrutiny.

**fMRI and PRS Data Analysis**

**Partial least squares analysis (PLS).** Partial least squares correlation (i.e., PLS, 45), a multivariate data-driven manner technique which can identify relationships between neural patterns (latent variables or LVs) and individual differences variables (behavioural PLS), was used to probe the transcriptomic and functional brain profiles linked to AD/MDD-related genetic vulnerability, psychological resilience and accelerated biological ageing. PLS was implemented using a series of Matlab scripts, which are available for download at https://www.rotman-baycrest.on.ca/index.php?section=345.

Two sets of behavioural PLS analyses were conducted. The sole analysis in the first set featured resilience, biological ageing, as well as the MDD and AD (APOE- vs non-APOE-based) PRSs in the "behavioural" matrix. The brain matrix contained the flexibility indices estimated in NCT and modelled as four separate conditions reflecting functional brain reorganisation, longitudinally within each task (incentive vs inhibitory control), as well as between the two tasks at baseline and the two-year follow-up, respectively.

The second set of PLS analyses sought to characterise the transcriptomic signature of resilience, biological ageing and AD/MDD genetic vulnerability. Specifically, it estimated the correlation between the brain LVs identified in behavioural PLS analysis 1 and the bi-hemispheric (analysis 1)/left-hemisphere (analysis 2) parcel x gene expression level matrix derived with the abagen toolbox from the comprehensive transcriptomic maps provided by the Allen Brain Institute.

***Significance and reliability testing.*** In all the reported PLS analyses, the significance of each LV was determined using a permutation test (5000 permutations for the brain-[behavior] analyses and 100,000 permutations for all the analyses involving gene expression data). In the permutation test, the rows of the parcel or of the gene expression data are randomly reordered (45). In all PLS analyses, potential axis rotations (i.e., changes in the order of the extracted LVs) and reflections (i.e., changes in the sign of the saliences), which may occur during resampling with either permutations or bootstrapping, were corrected with a Procrustes rotation, which defines a transformation through which the resampled singular value decomposition outcome (i.e., the identified LVs) is rotated to match most closely the original singular value decomposition outcome (46).

In the gene-brain PLS analysis, to account for correlated gene expression patterns based on anatomical proximity (47) we used Vasa's "rotate_parcellation" function in Matlab (https://github.com/frantisekvasa/rotate_parcellation/commit/bb8b0ef10980f162793cc180cef371e83655c505) in order to generate 100,000 spatially constrained permutations of the Schaefer brain LV, as identified in behavioral PLS analysis 1. These spatially constrained permuted brain LVs were used to assess the significance of the extracted gene LVs.

The reliability of each parcel's contribution to a particular LV was tested by submitting all weights to a bootstrap estimation (1000 bootstraps for the brain-[behaviour] analyses and 100000 bootstraps for all the analyses involving gene expression data) of the standard errors (SEs) (the bootstrap samples were obtained by sampling with replacement from the participants, 45). In order to increase the stability of the reported results, we used a number of permutations/bootstraps greater than the standard ones (i.e., 500 permutations/100 bootstrap samples), as recommended by [48] for use in PLS analyses of neuroimaging data. The higher number of permutations/bootstraps used for the gene expression data was determined by the relatively lower result stability compared to the brain-(behaviour) only analyses, as suggested by preliminary successive iterations of the gene PLS analysis using 5000 permutations/1000 bootstraps. A bootstrap ratio (BSR) (weight/SE) of at least 3 in absolute value (approximate associated p-value < .005) was used as a threshold for identifying those parcels that made a significant contribution to the identified LVs. For the gene PLS analyses, we focused on approximately the top 10% of absolute value BSRs (i.e., ~4, associated $p$-value < $10^{-4}$).

**Mediation analyses.** To test whether perinatal adversity and/or accelerated biological ageing modulate the neurodevelopmental correlates of resilience as a function of genetic risk for AD and MDD, we conducted two moderated mediation analyses using Hayes' PROCESS 3.5 macro for SPSS (49). These analyses were based on the PLS results and, as such, for AD, they only involved the no-APOE-based PRS. Two simple mediation analyses involving the same predictors, mediators and outcome were also performed for exploratory purposes. PROCESS is an ordinary least squares (OLS) and logistic regression path analysis modelling tool, based on observable variables. Moderated mediation models were tested employing 95% CI with 50000 bootstrapping samples. In line with extant guidelines on balancing Type I and Type II errors in mediation analyses (50), the CIs for indirect effects was estimated using percentile bootstrap, which is the default option in PROCESS 3.5. As recommended by [51], a heterodasticity consistent standard error and covariance matrix estimator was used. Bootstrapping-based 95% CIs for the indirect effects and for the moderation mediation index, as outputted by PROCESS, were used as effect size estimates.

**Replication of Results with the Gordon Atlas**

All the results described in the main report were replicated using data from a distinct functional brain atlas [52].

**Parcel definition.** Our main analyses based on the Schaefer atlas were replicated using the 333-node parcellation, proposed by [52] and downloaded from https://sites.wustl.edu/petersenschlaggarlab/resources/ This atlas was selected because it comprises all so-called core functional brain networks, which are also part of the Schaefer atlas, such as auditory (AUD), cingulo-opercular (CON), cingulo-parietal (CP), default mode (DMN), dorsal attention (DAN), frontoparietal (FPC), retrosplenial (RSP), sensorimotor-hand (SM-hand), sensorimotor-mouth (SM-mouth), salience (SAL), ventral attention (VAN) and visual (VIS). The replication analyses used only the 286 parcels which, based on Gordon et al.'s analyses, showed relatively unambiguous membership to one of the large-scale functional networks outlined above. We implemented this step in order to increase cross-atlas comparability since all the Schaefer parcels had a relatively unambiguous functional network assignment.

#### Mesoscale Functional Brain Correlates of Resilience and AD/MDD Risk

PLS analysis 1 identified two LVs (*p*-values of .0002 and .0004, respectively), which accounted for 18% and 12%, respectively, of the brain-behaviour covariance. The extracted brain LVs showed behavioural associations and functional flexibility patterns similar to the ones reported in the main text for the Schaefer atlas (see Figures S1-2).

#### Microscale Correlates of Resilience and AD/MDD Risk

##### MDD-Relevant Gene Expression Profiles.

***Gene-brain PLS: Left-hemisphere only.*** The gene-brain PLS analysis identified a sole LV ($p$ = .038), which accounted for 66% of the brain-gene covariance. The extracted gene LV was positively correlated both the brain LV1 (cf. Figure S1), $r$ = .39, 95% CI = [.33; .54] and brain LV2 (cf. Figure S2), $r$ = .43, 95% CI = [.36; .57].

***Gene-brain PLS: Bi-hemispheric.*** The above brain-gene pattern was replicated using the bi-hemispheric gene expression data from the Schaefer atlas ($p$ = .040, 69% brain-gene covariance explained; see Figure S7-a, b for the spatial expression profile of the gene LV).

As expected, the identified gene LV was robustly related to both brain LVs (LV1: $r$ = .27, 95% CI = [.20; .38]; LV2: $r$ = .45, 95% CI = [.41; .54]).

***MDD-relevant gene expression profile.*** Based on the brain-gene LV correlations, MDD-relevant transcriptomic associations with resilience, accelerated biological ageing and higher MDD PRS were derived from on the number of MDD_low genes with negative BSRs and MDD_high genes with positive BSRs. Complementarily, the number of the MDD_high genes with negative BSRs and MDD_low genes with positive BSRs was used to elucidate whether the neural profile associated with no-APOE-based AD risk (see Figure S2-a) would be typified by stronger expression of the MDD-relevant gene expression profile. No significant association was detected between the MDD-relevant gene expression and the resilience-linked brain LVs ($p$-values of .20 and .077 based on left hemisphere and bi-hemispheric data, respectively). In contrast, we found evidence of a link between the MDD-relevant transcriptomic profile and the neural profile linked to the no-APOE AD PRS ($p$-values of .006 and .068 based on left hemisphere and bi-hemispheric data, respectively; for relative MDD gene contribution, see Figure S7-c, d).

**Receptor Density Maps**

Using the same approach as the one described in the main text, we replicated the association between the brain LV1 and the DA receptor density map ($r$ of .19, spin test $p$ = .033), as well as the one between the brain LV2 and the E/I density map ($r$ of -.21, spin test $p$ = .045) (see Figure S8-a, d for the scatter plots describing these relationships and Figure S8-b, c, e for the E/I and DA receptor density maps).

**Moderated Mediation Analyses**

**Accelerated biological ageing strengthens the link between genetic risk for MDD and neurodevelopmental patterns predictive of subsequent resilience.** We replicated the partial moderated mediation analysis model reported in the main text, index of .008, SE =

.006, 90% CI [.0002; .020]. Specifically, we found that brain LV1 scores were predicted by a robust MDD PRS x biological ageing interaction, $b$ = .143, SE = .065, $t$(974) = 2.214, $p$ = .027 (see Figure S9-b), such that the link between the brain LV1 and MDD PRS was only significant for the fastest developing participants, effect of .141, SE = .045, 95% CI [.053; .230]. Accordingly, follow-up analyses confirmed that a significant mediation of MDD PRS effects on resilience via the brain LV1 was observed only for the fastest developing participants, effect of .008, SE = .005, 90% CI [.001; .018], biological ageing.

**Simple mediation models.** Two parallel mediator models replicated the findings observed with the Schaefer atlas: brain LV2 partially explained the indirect effect (IE) of no-APOE-based AD vulnerability on reduced resilience, IE = -.014, SE = .006, 95% CI [-.027; -.004] (see Figure S9-a), as well as the association between the MDD PRS and greater resilience, IE = .022, SE = .007, 95% CI [.009; .037] (see Figure 8-b).

18. Hagler DJ, Jr, Hatton, S., Cornejo, M.D., Makowski, C., Fair, D.A., Dick, A.S., Sutherland, M.T., Casey, B.J., Barch, D.M., Harms, M.P., Watts, R., Bjork, J.M., Garavan, H.P., Hilmer, L., Pung, C.J., Sicat, C.S., Kuperman, J., Bartsch, H., Xue, F., Heitzeg, M.M., Laird, A.R., Trinh, T.T., Gonzalez, R., Tapert, S.F., Riedel, M.C., Squeglia, L.M., Hyde, L.W., Rosenberg, M.D., Earl, E.A., Howlett, K.D., Baker, F.C., Soules, M., Diaz, J., de Leon, O.R., Thompson, W.K., Neale, M.C., Herting, M., Sowell, E.R., Alvarez, R.P., Hawes, S.W., Sanchez, W.K., Bodurka, J., Breslin, F.J., Morris, A.S., Paulus, M.P., Simmons, W.K.,

Polimeni, J.R., van der Kouwe, A., Nencka, A.S., Gray, K.M., Pierpaoli, C., Matochik, J.A., Noronha, A., Aklin, W.M., Conway, K., Glantz, M., Hoffman, E., Little, R., Lopez, M., Barch, V., Weiss, S.R., Wolff-Hughes, D.L., DelCarmen-Wiggins, R., Feldstein Ewing, S.W., MirandaDominguez, O., Nagel, B.J., Perrone, A.J., Sturgeon, D.T., Goldstone, A., Pfefferbaum, A., Pohl, K.M., Prouty, D., Uban, K., Bookheimer, S.Y., Dapretto, M., Galvan, A., Bagot, K., Giedd, J., Infante, M.A., Jacobus, J., Patrick, K., Shilling, P.D., Desikan, R., Li, Y., Sugrue, L., Banich, M.T., Friedman, N., Hewitt, J.K., Hopfer, C., Sakai, J., Tanabe, J., Cottler, L.B., Nixon, S.J., Chang, L., Cloak, C., Nagel, T., Reeves, G., Kennedy, D.N., Heeringa, S., Peltier, S., Schulenberg, J., Sripada, C., Zucker, R.A., Iacono, W.G., Luciana, M., Calabro, F.J., Clark, D.B., Lewis, D.A., Luna, B., Schirda, C., Brima, T., Foxe, J.J., Freedman, E.G., Mruzek, D.W., Mason, M. J., Huber, R., McGlade, E., Prescot, A., Renshaw, P.F., Yurgelun-Todd, D.A., Allgaier, N.A., Dumas, J.A., Ivanova, M., Potter, A., Florsheim, P., Larson, C., Lisdahl, K., Charness, M.E., Fuemmeler, B., Hettema, J.M., Maes, H.H., Florsheim, J., Anokhin, A.P., Glaser, P., Heath, A.C., Madden, P.A., Baskin-Sommers, A., Constable, R.T., Grant, S.J., Dowling, G.J., Brown, S.A., Jernigan, T.L., Dale, A.M., 2019. Image processing and analysis methods for the Adolescent Brain Cognitive Development Study. NeuroImage, 202, 116091.

19. Schaefer, A., Kong, R., Gordon, E. M., Laumann, T. O., Zuo, X. N., Holmes, A. J., Eickhoff, S. B., & Yeo, B. (2018). Local-Global Parcellation of the Human Cerebral Cortex from Intrinsic Functional Connectivity MRI. Cerebral Cortex, 28, 3095–3114.

20. Finc, K., Bonna, K., He, X., Lydon-Staley, D. M., Kühn, S., Duch, W., & Bassett, D. S. (2020). Dynamic reconfiguration of functional brain networks during working memory training. Nature Communications, 11, 2435.

21. Mucha, P. J., Richardson, T., Macon, K., Porter, M. A., & Onnela, J. P. (2010). Community structure in time-dependent, multiscale, and multiplex networks. Science, 328, 876–878.

22. Mattar, M. G., Cole, M. W., Thompson-Schill, S. L., & Bassett, D. S. (2015). A Functional Cartography of Cognitive Systems. PLoS Computational Biology, 11, e1004533.

23. Good, B. H., de Montjoye, Y. A., & Clauset, A. (2010). Performance of modularity maximization in practical contexts. Physical review. E, Statistical, nonlinear, and soft matter physics, 81(4 Pt 2), 046106.

24. Howard, D. M., Adams, M. J., Clarke, T. K., Hafferty, J. D., Gibson, J., Shirali, M. et al. (2019). Genome-wide meta-analysis of depression identifies 102 independent variants and highlights the importance of the prefrontal brain regions. Nature neuroscience, 22, 343–352.

25. Kunkle, B. W., Grenier-Boley, B., Sims, R., Bis, J. C., Damotte, V., Naj, A. C et al. (2019). Genetic meta-analysis of diagnosed Alzheimer's disease identifies new risk loci and implicates Aβ, tau, immunity and lipid processing. Nature genetics, 51, 414–430.

26. Watanabe, K., Taskesen, E., van Bochoven, A., & Posthuma, D. (2017). Functional mapping and annotation of genetic associations with FUMA. Nature Communications, 8, 1826.

27. Leonenko, G., Shoai, M., Bellou, E., Sims, R., Williams, J., Hardy, J., Escott-Price, V., & Alzheimer's Disease Neuroimaging Initiative (2019). Genetic risk for Alzheimer Disease is distinct from genetic risk for amyloid deposition. Annals of Neurology, 86, 427–435.

28. Fang, Y., Scott, L., Song, P., Burmeister, M., & Sen, S. (2020). Genomic prediction of depression risk and resilience under stress. Nature Human Behaviour, 4, 111–118.

29. Martens, Y. A., Zhao, N., Liu, C. C., Kanekiyo, T., Yang, A. J., Goate, A. M., Holtzman, D. M., & Bu, G. (2022). ApoE Cascade Hypothesis in the pathogenesis of

Alzheimer's disease and related dementias. Neuron, S0896-6273(22)00229-X. Advance online publication. https://doi.org/10.1016/j.neuron.2022.03.004

30. Chang, C. C., Chow, C. C., Tellier, L. C., Vattikuti, S., Purcell, S. M., & Lee, J. J. (2015). Second-generation PLINK: rising to the challenge of larger and richer datasets. GigaScience, 4, 7.

31. Choi, S. W., Mak, T. S., & O'Reilly, P. F. (2020). Tutorial: a guide to performing polygenic risk score analyses. Nature Protocols, 15, 2759–2772.

32. Markello, R. D., Arnatkeviciute, A., Poline, J. B., Fulcher, B. D., Fornito, A., & Misic, B. (2021). Standardizing workflows in imaging transcriptomics with the abagen toolbox. eLife, 10, e72129.

33. Arnatkevičiūtė, A., Fulcher, B. D. & Fornito, A. (2019) A practical guide to linking brain-wide gene expression and neuroimaging data. NeuroImage, 189, 353–367.

34. Hawrylycz, M., Miller, J. A., Menon, V., Feng, D., Dolbeare, T., Guillozet-Bongaarts, A. L., … & Lein, E. (2015). Canonical genetic signatures of the adult human brain. Nature Neuroscience, 18, 1832.

35. Fulcher, B. D., Little, M. A., & Jones, N. S. (2013). Highly comparative time-series analysis: the empirical structure of time series and their methods. Journal of the Royal Society Interface, 10, 20130048.

36. Petrican, R., Fornito, A., & Jones, N. (2022). Psychological resilience and neurodegenerative risk: A connectomics-transcriptomics investigation in healthy adolescent and middle-aged females. NeuroImage, 255, 119209.

37. Hansen, JY, Shafiei G, Markello, RD, Smart, K, Cox, SML, Wu, Y, Gallezot, J-D, Aumont, E, Servaes, S, Scala, SG, DuBois, JM, Wainstein, G, Bezgin, G, Funck, T, Schmitz, TW, Spreng, RN, Soucy, J-P, Baillet, S, Guimond, S, Hietala, J, Bédard, M-A, Leyton, M, Kobayashi, E, Rosa-Neto, P, Palomero-Gallagher, N, Shine, JM, Carson, RE, Tuominen, L,

Dagher A, & Misic B (2021). Mapping neurotransmitter systems to the structural and functional organization of the human neocortex. Biorxiv. doi:10.1101/2021.10.28.466336.

38. Prévot, T., & Sibille, E. (2021). Altered GABA-mediated information processing and cognitive dysfunctions in depression and other brain disorders. Molecular Psychiatry, 26, 151–167.

39. Johnson, D., Policelli, J., Li, M., Dharamsi, A., Hu, Q., Sheridan, M. A., McLaughlin, K. A., & Wade, M. (2021). Associations of Early-Life Threat and Deprivation With Executive Functioning in Childhood and Adolescence: A Systematic Review and Meta-analysis. JAMA pediatrics, 175, e212511.

40. Diemer, M.A., Mistry, R.S., Wadsworth, M.E., Lopez, I., & Reimers, F. (2012). Best practices in conceptualizing and measure social class in psycholgocial research. Analyses of Social Issues and Public Policy, 13, 77–113.

41. Moos, R.H. & Moos, B.S. (1994). Family Environment Scale Manual. Consulting Psychologists Press, Palo Alto, CA.

42. Echeverria, S.E., Diez-Roux, A.V., & Link, B.G. (2004). Reliability of self-reported neighborhood characteristics. Journal of Urban Health, 81, 682–701

43. Winkler, A. M., Renaud, O., Smith, S. M., & Nichols, T. E. (2020). Permutation inference for canonical correlation analysis. NeuroImage, 220, 117065.

44. Hair, J. F., Black, W. C., Babin, B. J., Anderson, R. E., & Tatham, R. L. (2014). Multivariate data analysis. Pearson Education Limited.

45. Krishnan, A., Williams, L. J., McIntosh, A. R., & Abdi, H. (2011). Partial Least Squares (PLS) methods for neuroimaging: a tutorial and review. NeuroImage, 56, 455–475.

46. Milan, L. & Whittaker, J. (1995). Application of the parametric bootstrap to models that incorporation a singular value decomposition. Journal of the Royal Statistical Society, Series C (Applied Statistics), 44, 31–49.

47. Fornito, A., Arnatkevicuite, A., Fulcher, B. D. (2019). Bridging the gap between the brain's transcriptome and connectome. Trends in Cognitive Sciences, 23, 34-50.

48. McIntosh, A. R., & Lobaugh, N. J. (2004). Partial least squares analysis of neuroimaging data: applications and advances. NeuroImage, 23 Suppl 1, S250–S263.

49. Hayes, A.F. (2018) Introduction to Mediation, Moderation, and Conditional Process Analysis: A Regression-Based Approach. (Second Edition) Guilford Press: New York.

50. Hayes, A.F. & Scharkow, M. (2013) The relative trustworthiness of inferential tests of the indirect effect in statistical mediation analysis: does method really matter? Psychological Science, 10, 1918-1927.

51. Hayes, A. & Cai, L. (2007) Using heteroskedasticity-consistent standard error estimators in OLS regression: An introduction and software implementation. Behavior Research Methods, 39, 709-722.

52. Gordon, E.M., Laumann, T.O., Adeyemo, B., Huckins, J.F., Kelley, W.M., & Petersen, S.E. (2016) Generation and evaluation of a cortical area parcellation from resting-state correlations. Cerebral Cortex, 26, 288-303.

Table S1

*MNI Coordinates, Anatomical Location and Functional Network Allegiance for the Schaefer and Gordon Parcels with No Reliable Gene Expression Data*

| | MNI Coordinates | | | Anatomical Location | Functional Network Allegiance |
|---|---|---|---|---|---|
| Schaefer atlas | X | Y | Z | | |
| 1 | -50 | -12 | 14 | Primary sensory cortex | SM- B |
| 2 | -50 | -16 | 44 | Primary sensory cortex | SM-B |
| 3 | -12 | -4 | 72 | BA6 | SAL-VAN |
| Gordon atlas | | | | | |
| 1 | -27.5 | -37.2 | 61.4 | Sensory association cortex | SM-H |
| 2 | -44.8 | -54 | 14.6 | Angular gyrus | VAN |
| 3 | -46.3 | -41.4 | 25.9 | Angular gyrus | AUD |
| 4 | -45.4 | 28.8 | 0.8 | BA45 | VAN |
| 5 | -52.2 | -14.1 | 15.2 | Primary sensory cortex | AUD |
| 6 | -34.7 | 5.6 | 34 | Frontal eye fields | DAN |

| | | | | | |
|---|---|---|---|---|---|
| 7 | -41.6 | 8.7 | 22.2 | BA44 | DAN |
| 8 | -27.5 | 53.6 | 0 | BA10 | DMN |
| 9 | -26.6 | 46.8 | 20.9 | BA10 | CON |
| 10 | -35.7 | 33.1 | 32 | BA9 | DAN |
| 11 | -38.7 | 4.8 | 48.4 | BA6 | VAN |
| 12 | 6.7 | 5 | 55.9 | BA6 | CON |
| 13 | 16.2 | 0.8 | 67.5 | BA6 | CON |
| 14 | 28 | -34.8 | 63.1 | Primary sensory cortex | SM-H |
| 15 | 30.6 | 22.8 | -4.7 | Insula | SAL |
| 16 | 26.8 | -55 | 54.2 | Visuomotor cortex | VIS |
| 17 | 7.7 | -85.6 | 31.6 | Visual association cortex | VIS |
| 18 | 54.2 | -13.6 | 16.9 | Supramarginal gyrus | AUD |
| 19 | 31.2 | -45.6 | -5.8 | Visual association cortex | VIS |
| 20 | 20.4 | -87.3 | -6.6 | Secondary visual cortex | VIS |

Note. BA = Brodmann area. Schaefer networks: SAL-VAN = salience/ventral attention. SM-B =somatomotor-B. Gordon networks: AUD = auditory. CON = cingulo-opercular. DMN = default mode. DAN = dorsal attention. VAN = ventral attention. SM-H = somatomotor-hand. VIS = visual.

#### Supplementary Figure Captions

*Figure S1*. First extracted LV from the behavioral-PLS analysis linking AD/MDD genetic risk, resilience, and biological ageing to longitudinal and contemporaneous cross-context differentiation of the functional brain architecture relevant to incentive processing vs inhibitory control. Panel (a) shows the correlations between the behavioral variables and the brain scores in each condition. Panel (b) depicts the Gordon ROIs with robust loadings (absolute value BSR > 3) on the LV in panel (a) and visualized with the BrainNet Viewer (http://www.nitrc.org/projects/bnv/) (Xia et al., 2013). ROI colours reflect Gordon et al.'s network assignments. In panel (b), the size of the ROIs is proportional to their associated absolute value BSR. Error bars are the 95% confidence intervals from the bootstrap procedure. Confidence intervals that do not include zero reflect robust correlations between the respective behavioral variable and the brain score in a given condition across all participants. In panel (a), hypothesis-relevant, significant brain-behavior correlations replicating the main analyses, are within red line rectangles. Significant brain-behavior correlations unrelated to our hypotheses are in blue rectangles. LV= latent variable. PRS = polygenic risk score. BSR = bootstrap ratio. Gordon networks: AUD = auditory; CON = cingulo-opercular; CP = cingulo-parietal; DMN = default mode; DAN = dorsal attention; FPC = frontoparietal; RSP = retrosplenial/temporal; SM-H =somatomotor-hand; SM-mouth = somatomotor mouth; SAL = salience; VAN = ventral attention; VIS =visual.

*Figure S2*. Second extracted LV from the behavioral-PLS analysis linking AD/MDD genetic risk, resilience, and biological ageing to longitudinal and contemporaneous cross-context differentiation of the functional brain architecture relevant to incentive processing vs inhibitory control. Panel (a) shows the correlations between the behavioral variables and the brain scores in each condition. Panels (b) and (c) depict the Gordon ROIs with robust loadings (absolute value BSR > 3) on the LV in panel (a) and visualized with the BrainNet

Viewer (http://www.nitrc.org/projects/bnv/) (Xia et al., 2013). ROI colours reflect Gordon et al.'s network assignments. In panel (b), the size of the ROIs is proportional to their associated absolute value BSR. Error bars are the 95% confidence intervals from the bootstrap procedure. Confidence intervals that do not include zero reflect robust correlations between the respective behavioral variable and the brain score in a given condition across all participants. In panel (a), hypothesis-relevant, significant brain-behavior correlations replicating the main analyses, are within red line rectangles. Significant brain-behavior correlations unrelated to our hypotheses are in blue rectangles. LV= latent variable. PRS = polygenic risk score. BSR = bootstrap ratio. Gordon networks: AUD = auditory; CON = cingulo-opercular; CP = cingulo-parietal; DMN = default mode; DAN = dorsal attention; FPC = frontoparietal; RSP = retrosplenial/temporal; SM-H =somatomotor-hand; SM-mouth = somatomotor mouth; SAL = salience; VAN = ventral attention; VIS =visual.

*Figure S3.* Replication of the LV1-linked pattern (PLS analysis 1, see Figure 2-a), based on the Schaefer atlas data and using a variety of *p*-thresholds ($10^{-5}$ , $10^{-4}$ , $10^{-3}$ , $10^{-2}$) for selecting the AD/MDD PRS-contributing variants. For easier visualisation, we represented only the relevant PLS conditions for the *p*-thresholds (from the list above) at which our reported results had been broadly replicated. LV= latent variable. PRS = polygenic risk score. AD = Alzheimer's Disease. MDD = Major Depressive Disorder.

*Figure S4.* Replication of the LV1-linked pattern (PLS analysis 1b, see Figure S1-a), based on the Gordon atlas data and using a variety of *p*-thresholds ($10^{-5}$, $10^{-4}$ , $10^{-3}$ , $10^{-2}$) for selecting the AD/MDD PRS-contributing variants. For easier visualisation, we represented only the relevant PLS conditions for the *p*-thresholds (from the list above) at which our reported results had been broadly replicated. LV= latent variable. PRS = polygenic risk score. AD = Alzheimer's Disease. MDD = Major Depressive Disorder.

*Figure S5.* Replication of the LV2-linked pattern (PLS analysis 1, see Figure 3-a), based on the Schaefer atlas data and using a variety of *p*-thresholds ($10^{-5}$, $10^{-4}$, $10^{-3}$, $10^{-2}$) for selecting the AD/MDD PRS-contributing variants. For easier visualisation, we represented only the relevant PLS conditions for the *p*-thresholds (from the list above) at which our reported results had been broadly replicated. LV= latent variable. PRS = polygenic risk score. AD = Alzheimer's Disease. MDD = Major Depressive Disorder.

*Figure S6.* Replication of the LV2-linked pattern (PLS analysis 1, see Figure S2-a), based on the Gordon atlas data and using a variety of *p*-thresholds ($10^{-5}$, $10^{-4}$, $10^{-3}$, $10^{-2}$) for selecting the AD/MDD PRS-contributing variants. For easier visualisation, we represented only the relevant PLS conditions for the *p*-thresholds (from the list above) at which our reported results had been broadly replicated. LV= latent variable. PRS = polygenic risk score. AD = Alzheimer's Disease. MDD = Major Depressive Disorder.

*Figure S7.* Results of the MDD overlap analyses. Panels (a) and (b) represent the spatial expression map of the gene LV identified in the gene-brain behavioral PLS analysis. The ROIs were visualized with the BrainNet Viewer (http://www.nitrc.org/projects/bnv/) (Xia et al., 2013). ROI colours reflect Gordon et al.'s network assignments and their size is proportional to how strongly they express the gene LV (i.e., the absolute value of the associated brain score). Panels (c) and (d) depict the results of the MDD/non-APOE AD PRS overlap analyses, using only left-hemisphere (c) or bi-hemispheric (d) data. As described in the text, these are based on the gene LV depicted in panels (a) and (b), which was negatively correlated with the non-APOE PRS (see Figure S2-a). LV= latent variable. BSR = bootstrap ratio. Gordon networks: AUD = auditory; CON = cingulo-opercular; CP = cingulo-parietal; DMN = default mode; DAN = dorsal attention; FPC = frontoparietal; RSP = retrosplenial/temporal; SM-H =somatomotor-hand; SM-mouth = somatomotor mouth; SAL = salience; VAN = ventral attention; VIS =visual.

*Figure S8*. Results of the correlational analyses replicating the link of the E/I and DA receptor density map with the two brain LVs relevant to AD/MDD genetic risk, resilience and biological ageing (see Figures 2 and 3). Panels (a) and (d) contain the scatter plots describing the linear relationship between brain LV2 and the E/I (GLU/GABA) density map (a), as well as between brain LV1 and the DA density map (d). Panels (b) and (c) represent the areas of E > I density and I > E density, respectively, whereas panel (d) depicts the DA receptor density map. The ROIs were visualized with the BrainNet Viewer (http://www.nitrc.org/projects/bnv/) (Xia et al., 2013). ROI colours reflect Gordon et al.'s network assignments and their size is proportional to the density of the respective receptor. E/I = excitation/inhibition. GLU = glutamate. GABA = gamma-aminobutyric acid. DA = dopamine. AUD = auditory; CON = cingulo-opercular; CP = cingulo-parietal; DMN = default mode; DAN = dorsal attention; FPC = frontoparietal; RSP = retrosplenial/temporal; SM-H =somatomotor-hand; SM-mouth = somatomotor mouth; SAL = salience; VAN = ventral attention; VIS =visual.

*Figure S9*. (Moderated) mediational models linking genetic vulnerability to AD (panel a) and MDD (panel b), respectively to psychological resilience as assessed at the three-year follow-up. In both panels, the coefficients associated with the red paths had been derived from simple mediational analyses (as described in the text). The coefficients associated with the blue paths are based on the moderated mediation analyses. A dashed line indicates a statistically non-significant path ($p > .10$ [the one-sided $p < .05$ was considered appropriate for replication purposes).

### Mesoscale Functional Brain Markers

#### PLS 1b: Brain LV1

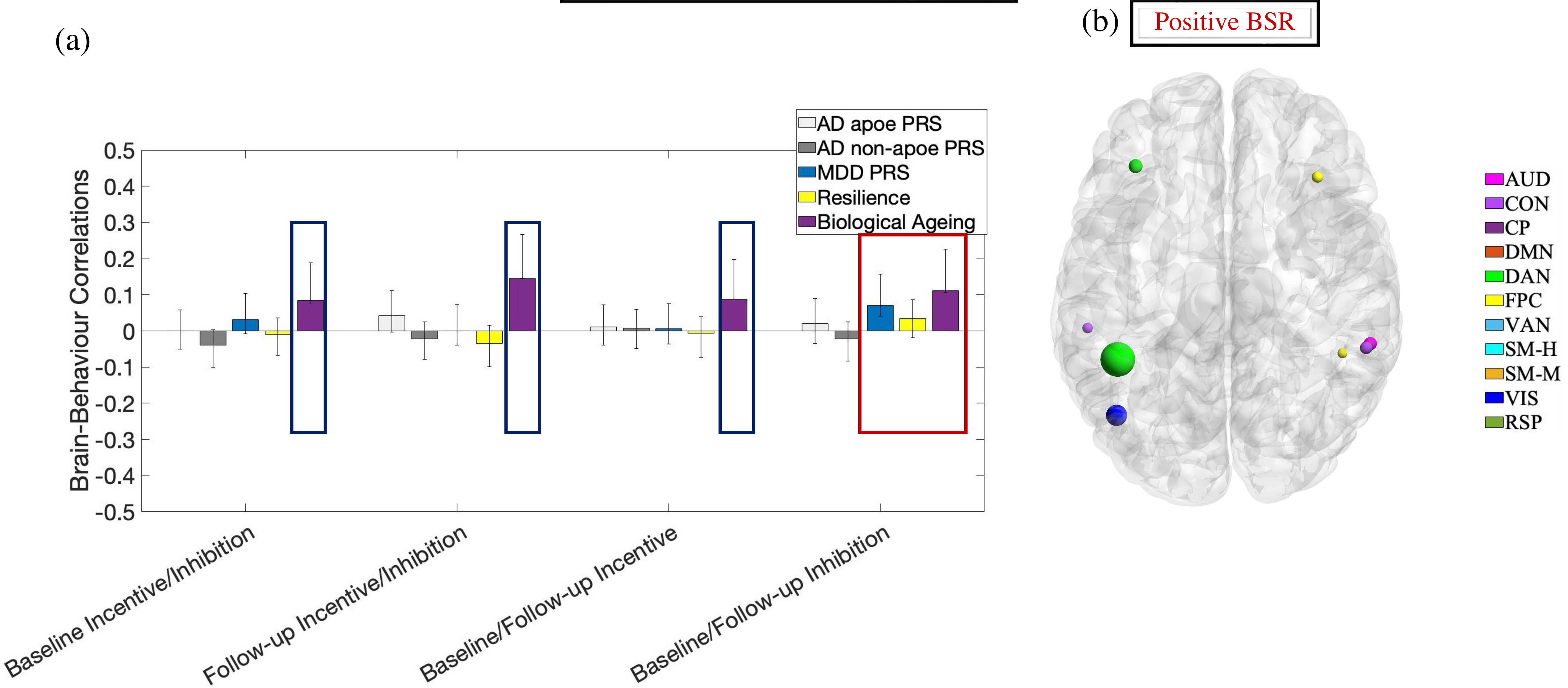


**Figure S1.**

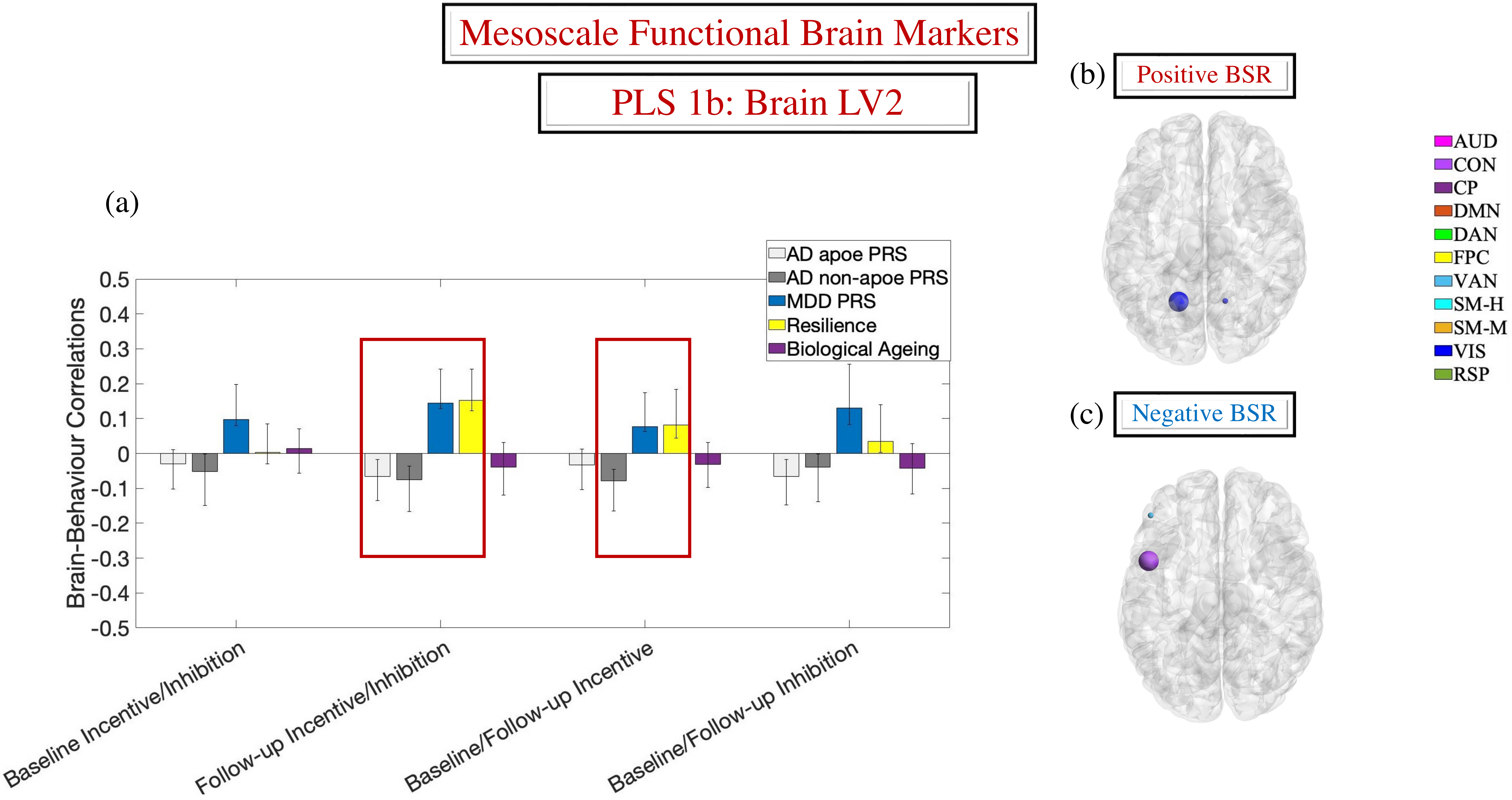
Mesoscale Functional Brain Markers
PLS 1b: Brain LV2
(a)
AD apoe PRS
AD non-apoe PRS
MDD PRS
Resilience
Biological Ageing
Brain-Behaviour Correlations
0.5
0.4
0.3
0.2
0.1
0
-0.1
-0.2
-0.3
-0.4
-0.5
Baseline Incentive/Inhibition
Follow-up Incentive/Inhibition
Baseline/Follow-up Incentive
Baseline/Follow-up Inhibition
(b)
Positive BSR
(c)
Negative BSR
AUD
CON
CP
DMN
DAN
FPC
VAN
SM-H
SM-M
VIS
RSP

**Figure S2.**

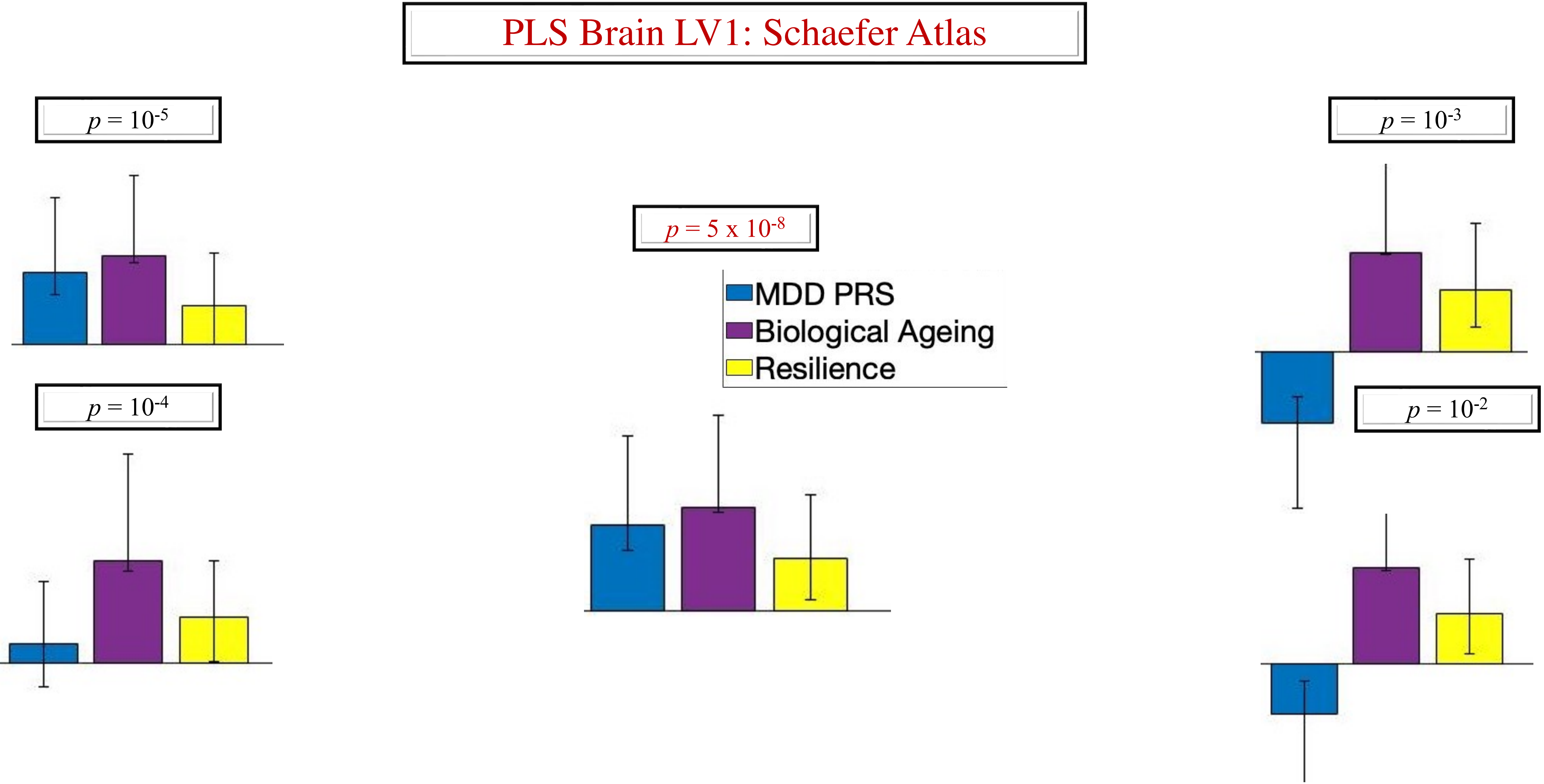
PLS Brain LV1: Schaefer Atlas
p = 10-5
p = 10-4
p = 5 x 10-8
MDD PRS
Biological Ageing
Resilience
p = 10-3
p = 10-2

**Figure S3.**

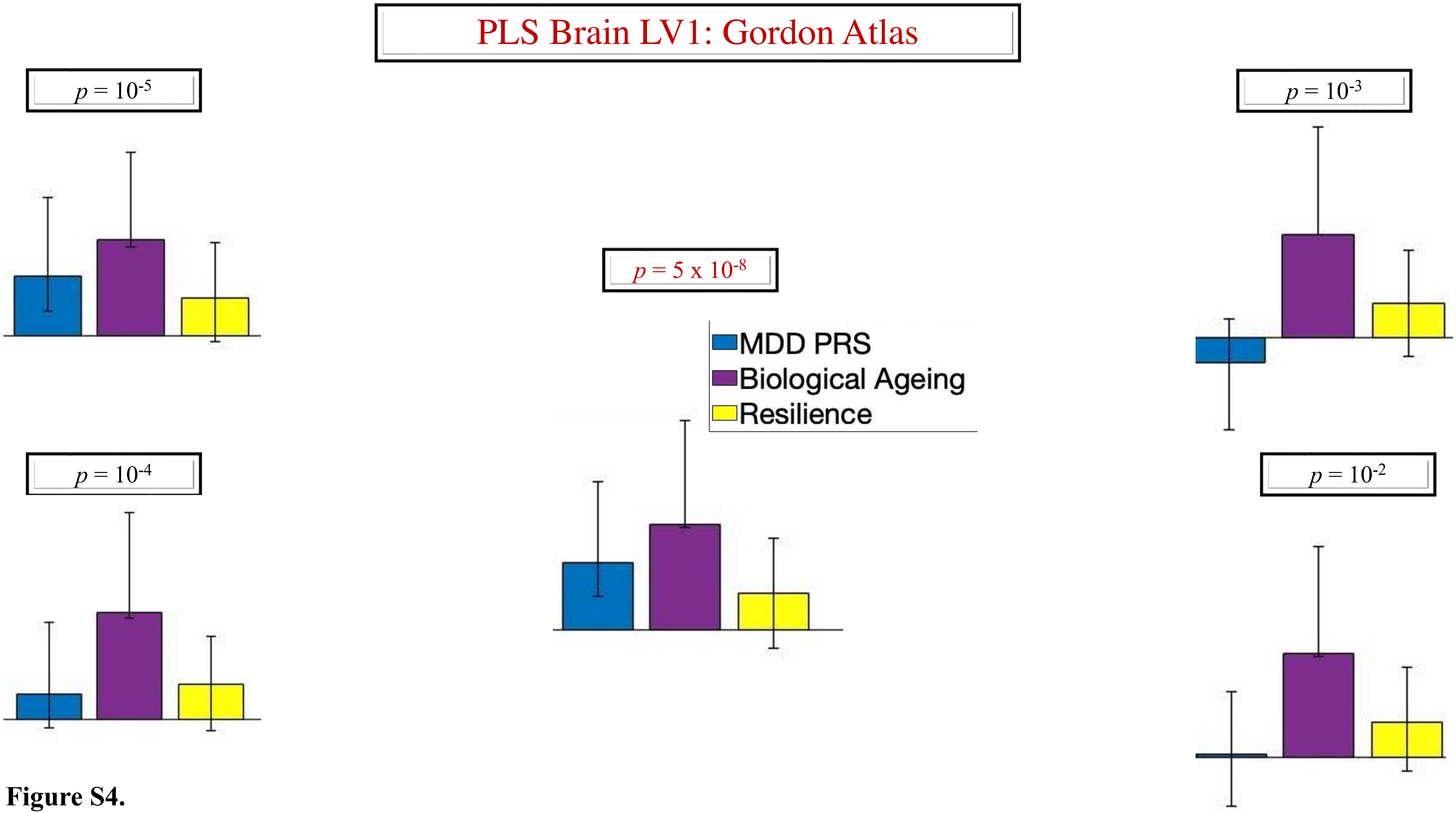
PLS Brain LV1: Gordon Atlas
$p = 10^{-5}$
$p = 10^{-3}$
$p = 5 \times 10^{-8}$
MDD PRS
Biological Ageing
Resilience
$p = 10^{-4}$
$p = 10^{-2}$

**Figure S4.**

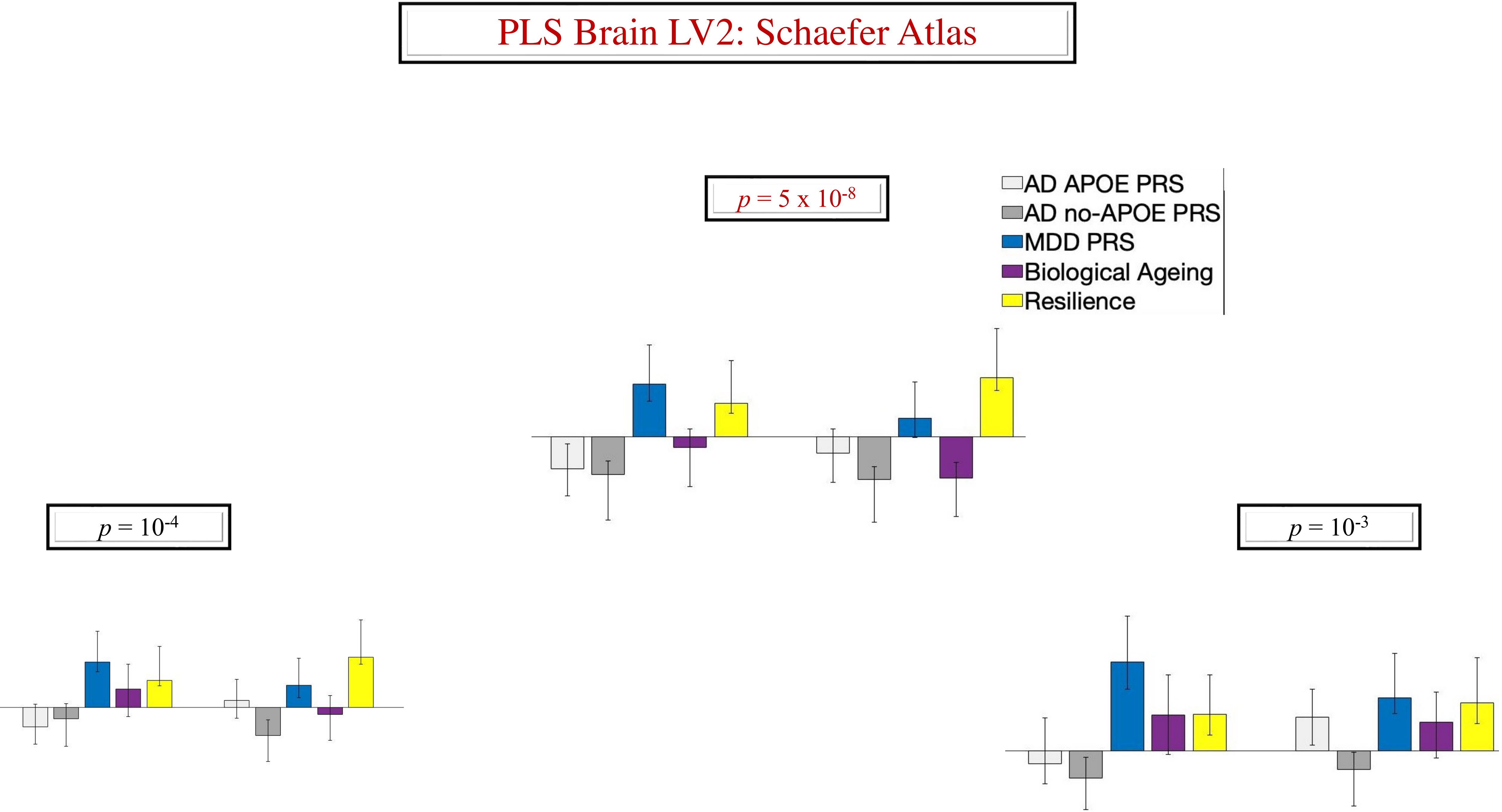
PLS Brain LV2: Schaefer Atlas
p = 5 x 10-8
AD APOE PRS
AD no-APOE PRS
MDD PRS
Biological Ageing
Resilience
p = 10-4
p = 10-3

**Figure S5.**

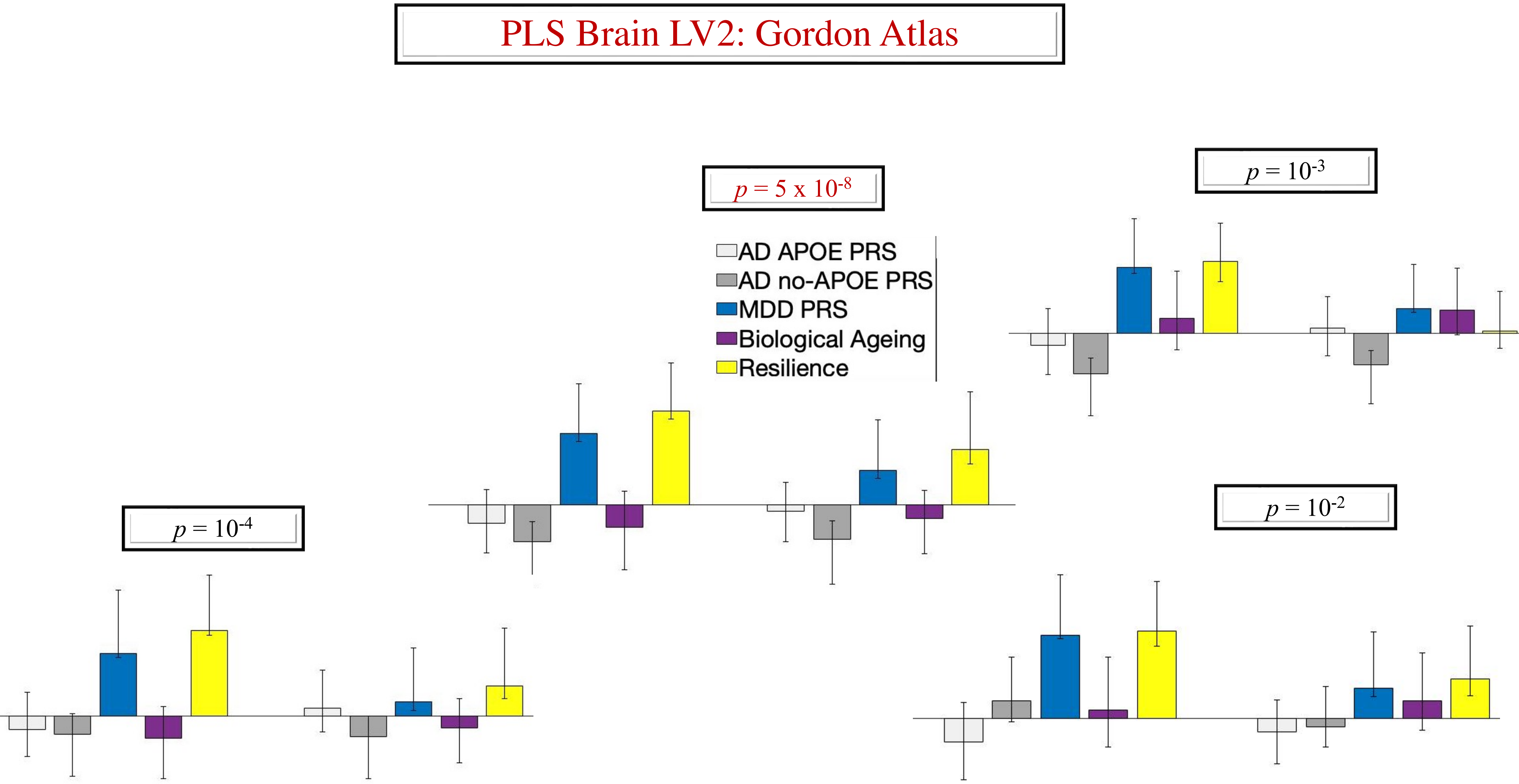
PLS Brain LV2: Gordon Atlas
p = 5 x 10-8
p = 10-3
AD APOE PRS
AD no-APOE PRS
MDD PRS
Biological Ageing
Resilience
p = 10-4
p = 10-2

**Figure S6.**

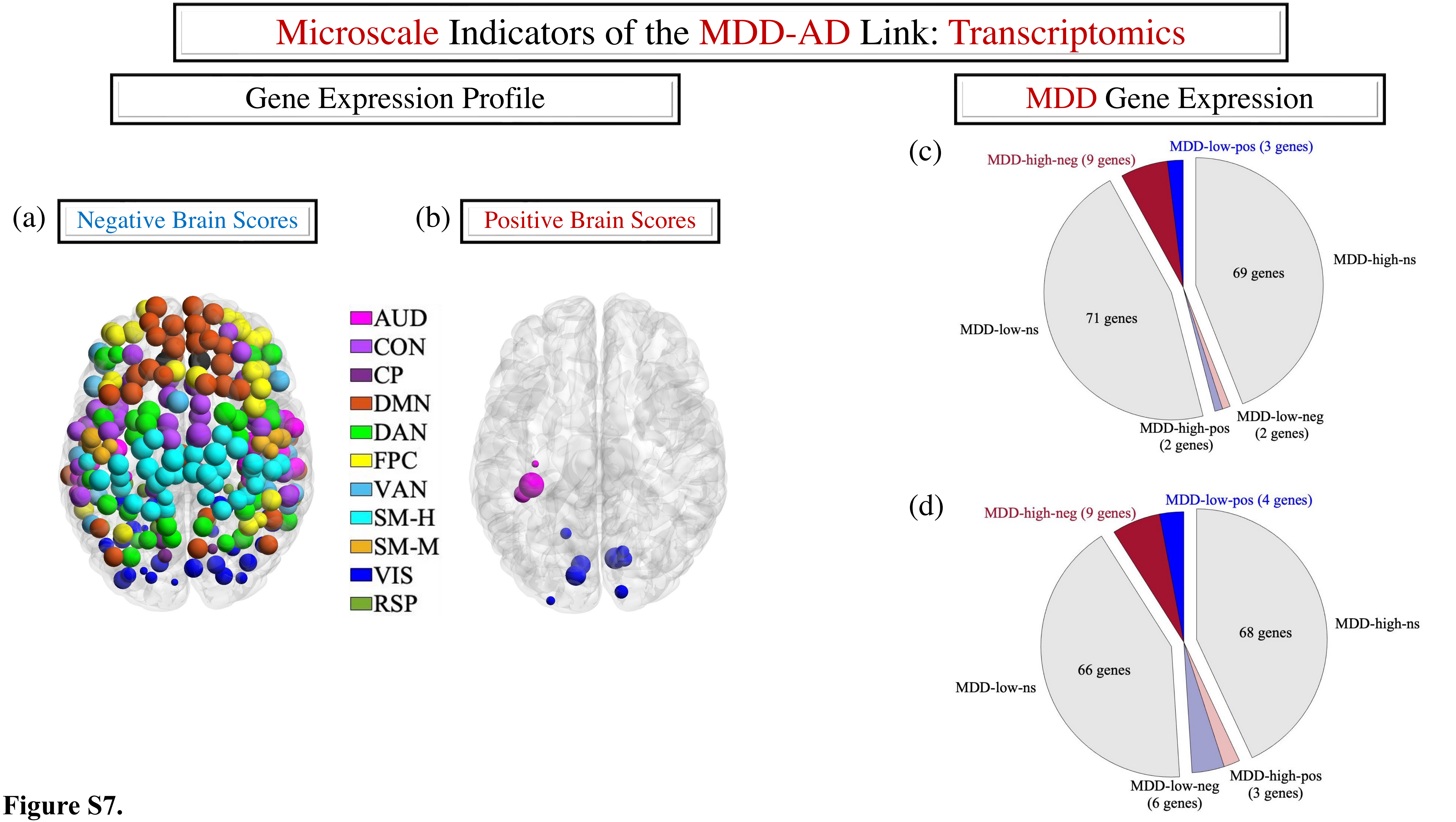

Microscale Indicators of the MDD-AD Link: Transcriptomics
Gene Expression Profile
MDD Gene Expression
(a)
Negative Brain Scores
(b)
Positive Brain Scores
AUD
CON
CP
DMN
DAN
FPC
VAN
SM-H
SM-M
VIS
RSP
(c)
MDD-high-neg (9 genes)
MDD-low-pos (3 genes)
MDD-high-ns
69 genes
71 genes
MDD-low-ns
MDD-high-pos (2 genes)
MDD-low-neg (2 genes)
(d)
MDD-high-neg (9 genes)
MDD-low-pos (4 genes)
MDD-high-ns
68 genes
66 genes
MDD-low-ns
MDD-low-neg (6 genes)
MDD-high-pos (3 genes)


**Figure S7.**

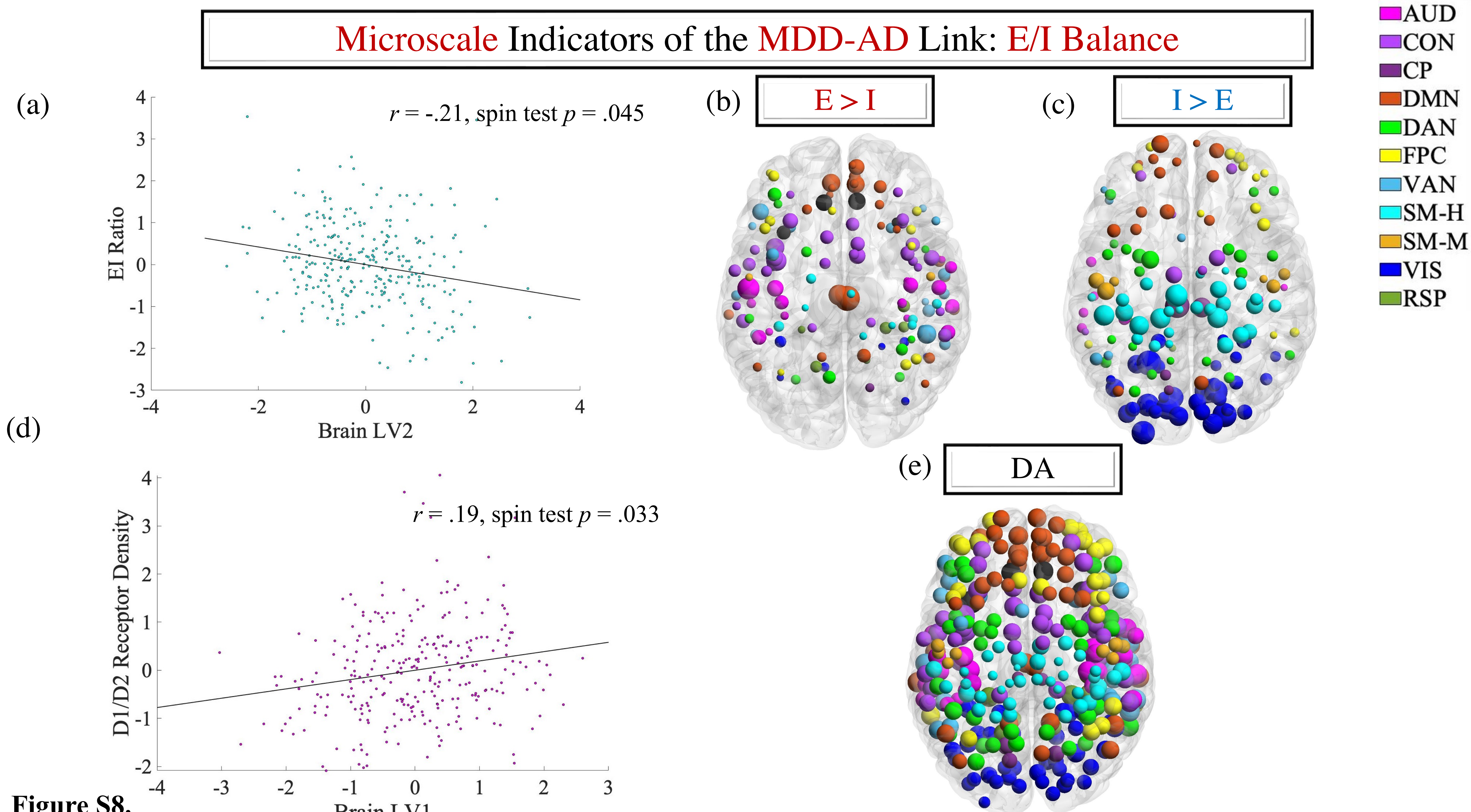

Microscale Indicators of the MDD-AD Link: E/I Balance
(a)
r = -.21, spin test p = .045
EI Ratio
Brain LV2
(b)
E > I
(c)
I > E
AUD
CON
CP
DMN
DAN
FPC
VAN
SM-H
SM-M
VIS
RSP
(d)
r = .19, spin test p = .033
D1/D2 Receptor Density
Brain LV1
(e)
DA


**Figure S8.**

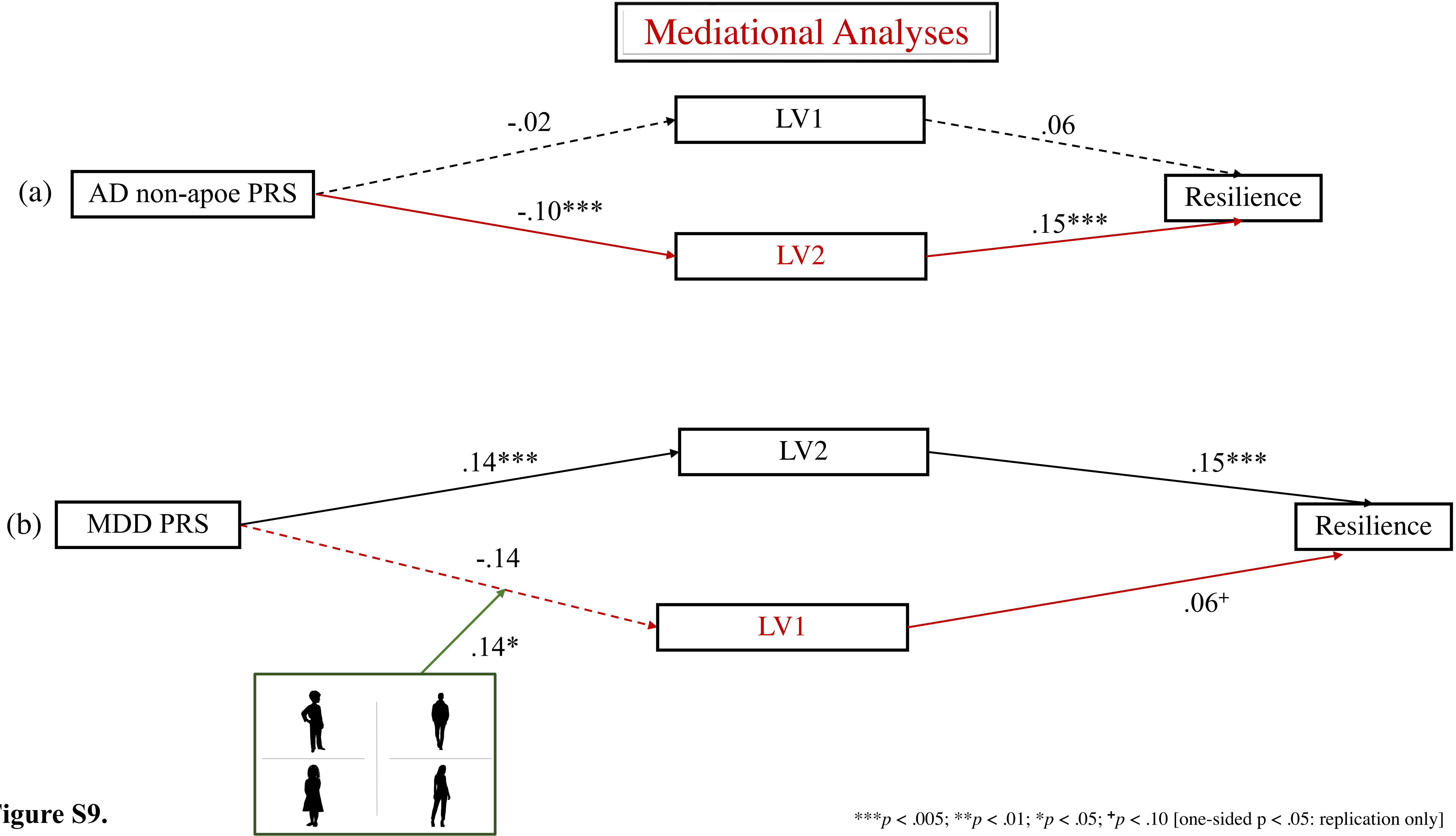


**Figure S9.**

***$p$ < .005; **$p$ < .01; *$p$ < .05; +$p$ < .10 [one-sided p < .05: replication only]
